# Durability of three types of dual active ingredient long-lasting insecticidal net compared to a pyrethroid-only LLIN in Tanzania: protocol for a prospective cohort study nested in a cluster randomized controlled trial

**DOI:** 10.1101/2021.04.25.21255903

**Authors:** Jackline L Martin, Louisa A. Messenger, Franklin W Mosha, Eliud Lukole, Jacklin F Mosha, Manisha Kulkarni, Thomas S Churcher, Ellie Sherrard-Smith, Alphaxard Manjurano, Natacha Protopopoff, Mark Rowland

## Abstract

**Introduction:** Progress achieved by long-lasting insecticidal nets (LLINs) against malaria is threatened by the widespread selection of pyrethroid resistance among vector populations. LLINs with non-pyrethroid insecticides are urgently needed. This study aims to assess the durability of three novel dual active ingredient LLINs and to parameterise a mathematical model to predict epidemiological outcomes of these products for malaria vector control.

**Methods:** A WHO Phase 3 active ingredients and textile durability study will be carried out within a cluster randomised controlled trial in Misungwi district in 40 clusters. The following treatments will be evaluated; 1/ Interceptor G2® combining chlorfenapyr and a pyrethroid alpha-cypermethrin, 2/ Royal Guard® treated with pyriproxyfen and alpha-cypermethrin and 3/ Olyset™ Plus which incorporates a synergist piperonyl butoxide and the pyrethroid permethrin, and 4/ a reference standard pyrethroid-only LLIN (Interceptor®). 750 nets will be followed in 5 clusters per intervention arm at 6, 12, 24 and 36 months post distribution for survivorship and hole index assessment. A second cohort of 1950 nets per each type will be identified in 10 clusters, of which 30 LLINs will be withdrawn for bio-efficacy and chemical analysis every 6 months up to 36 months and another 30 collected for an experimental hut trial study every year. Bio-efficacy will be assessed using cone bioassays and tunnel tests against susceptible and resistant laboratory strains of *Anopheles gambiae* sensu stricto. Efficacy of field collected nets will be compared in six experimental huts. The main outcomes will be *Anopheles* mortality up to 72 hours post exposure, blood feeding and egg maturation using ovary dissection to assess impact on fecundity.

**Ethics and dissemination:** Ethical approval was received from the Tanzanian ethics review committee as well as from each institution. Study findings will be disseminated via reports and presentations to national and international stakeholders, conferences, and peer-reviewed publications.

**Trial registration number:** NCT03554616

**Article summary:** *Strengths of this study:*

- This is the first study assessing the durability of these novel dual-A.I. LLINs over three years alongside a cluster randomised controlled trial (cRCT).
- The study will support the development of bio-efficacy and physical durability criteria of performance for the partner A.I. in relation to the cRCT efficacy outcomes and refine preferred product characteristics developed by the WHO.
- An experimental hut trial done in the vicinity of the cRCT, with similar vector population characteristics and using nets sampled from the main trial will allow us to understand the impact of field conditions, wear-and-tear and insecticidal deterioration on the efficacy of the dual-A.I. LLINs on entomological outcomes and relate these to the cRCT epidemiological and entomological outcomes.
- All other products falling under these novel dual-A.I. LLINs classes will be assessed according to those criteria alongside non-inferiority studies in experimental huts and modelling of these data.

*Limitations of this study:* Potential limitations include 1/ the net loss of follow up used to calculate sample size, which was taken from another study site and might differ in Misungwi, and 2/ the experimental hut site is situated in the northern section of the cRCT study area and may not represent vector species composition of the southern section of the cRCT.

## INTRODUCTION

Long-lasting insecticidal nets (LLINs) are the primary method of malaria control in sub-Saharan Africa. The World Health Organization (WHO) estimates that over 50% of the population now sleeps under LLINs. This has helped to reduce malaria incidence by 42% and mortality by 66% in Africa over the last 15 years.^1^ Until recently, pyrethroids were the only type of insecticide used routinely on LLINs. The rapid spread of pyrethroid resistance in vector populations threatens to reverse the success achieved so far.^2^ Several studies have demonstrated that LLINs are becoming less effective at killing mosquitoes in areas of high resistance compared to before.^3 4^

The first new type of LLIN developed to control resistant mosquitoes is a combination LLIN containing permethrin and the synergist piperonyl butoxide (PBO), which inhibits cytochrome P450 oxidases responsible for metabolic resistance. A community-based cluster randomized controlled trial (cRCT) conducted in Muleba demonstrated a reduction in the prevalence of malaria by 44% in the pyrethroid-PBO LLIN arm (Olyset™ Plus) compared to the standard pyrethroid LLIN after one year and by 33% after two years.^5^ Based on this study, WHO recognized the improved public health value of the pyrethroid-PBO LLIN in areas of high resistance and provided it an interim recommendation as a new class of vector control product.^6^ Since then, two other dual-active ingredient (dual-A.I.) LLINs (Royal Guard® and Interceptor G2®) have been evaluated in WHO Phase I and II trials and have shown promise compared to standard LLINs against pyrethroid resistant vectors. Each of these putative first-in-class LLINs are required by WHO to undergo cRCTs versus standard LLINs to demonstrate, unequivocally, evidence of improved malaria control effect.^7^ Two such community cRCTs are currently underway in Misungwi, Tanzania^8^ and in Cove, Benin.^9^

Assessment of the quality, efficacy and safety is also required for each vector control product submitted to WHO. Whether the LLIN product is a novel first-in-class LLIN, or a generic next-in-class LLIN, the LLIN should be evaluated in three different phases (Table 1).^10^

**Table 1:** Main parameters assessed in Phase I, II and III studies of LLINs.

| Phase | Type of study | Parameters measured |
| --- | --- | --- |
| I | Laboratory | 1. Regeneration of insecticidal activity<br>2. Efficacy and wash-resistance |
| II | Small-scale field trial | 1. Wash-resistance<br>2. Efficacy as measured by vector mortality and blood-feeding inhibition |
| III | Large-scale field trial | 1. Long-lasting insecticidal efficacy<br>2. Rate of loss or attrition of nets<br>3. Physical durability of netting material<br>4. Community acceptance<br>5. Safety |

In large-scale field trials, physical durability is affected by net care and repair, frequency of use, and maintenance practices, duration of transmission season, as well as textile physical features such as fibre material, knitting or weaving pattern.^11^ The WHO assumes a good LLIN will demonstrate a physical life span of 3 years but this duration can vary between product, endemic regions and condition of use.^12 13^ In the WHO guidelines, LLIN survivorship and fabric integrity are monitored in the community at 6, 12, 24 and 36 months.^10^ The pyrethroid component is expected to remain effective for 3 years,^10^ while the residual efficacy of other active ingredients are not yet known. ^12 13^

After a LLIN has been assessed for its durability (insecticidal and physical) in the field, the bio-efficacy of these nets can be estimated in experimental hut trials.^14 15^ Experimental huts are designed to mimic commonly used houses in the local area but modified to restrict the escape of mosquitoes and exclude scavenging insects; live and dead mosquitoes can be collected in the morning to assess insecticide-induced exiting, mortality and blood feeding inhibition. Currently, there are three designs of experimental hut recommended to evaluate LLINs and Indoor Residual Spraying (IRS) formulations.^10^ These are East African-style huts, West African-style huts and Asian style huts. During standard, prospective WHO Phase II evaluations of LLIN efficacy, each net is expected to retain insecticidal activity for at least 20 standardized washes with respect to vector knock-down, mortality and blood feeding inhibition.

Although community cRCTs provide definitive evidence for the establishment of new product classes of LLIN, questions remain whether cRCTs should be the sole mechanism to generate this evidence.^16^ It has been proposed that entomological evidence generated by experimental hut trials may be adequate to contribute to this judgement.^2^ The cost of cRCTs can be comparatively high, making returns on investment more challenging to achieve and this may act to disincentivise investments in new active ingredients and the criteria set out by the WHO, requiring two community-based cRCTs in two locations,^17^ may not be feasible for every new LLIN intervention class in a timely manner.^18^ Such factors may limit the number of available, approved malaria vector control 2^nd^ in class products for resistance management, allowing transmission to continue at present in areas of insecticide resistance.^18^

Some malaria transmission models make use of LLIN Phase II experimental hut trial data to estimate the impact of these nets on malaria incidence/prevalence over time.^2^ The WHO and advisory committees have ruled that the evidence base is presently insufficient to judge new classes of LLINs solely on the basis of entomological evidence.^19^

Malaria transmission dynamics mathematical models can be used to predict the public health impact of different vector control interventions.^20^ These models simulate infection in populations of humans and mosquitoes and could be used to extrapolate the results of cRCTs to other sites with different epidemiology, entomology or mixtures of control interventions. It is unclear whether models parameterised with local hut trial data capturing the behaviour of the local mosquito vector would be more accurate at predicting local epidemiology than this meta-analysis approach which uses data from disparate locations (with different mosquito populations). These models have typically used data from LLINs that have been subjected to standardised washing to simulate the aging process. As a result, their predicted ability may be limited by the accuracy of standardised washing to reflect real life wear-and-tear and insecticidal deterioration under field conditions.

The aims of this study are to assess the insecticidal and physical durability of new dual-A.I. LLINs, Interceptor G2®, Olyset™ Plus and Royal Guard® compared to a standard LLIN (Interceptor®) in the community in order to set durability criteria for these first in class dual-A.I. LLINs and to establish whether entomological outcomes generated during the WHO product evaluation process (adapted experimental hut trials and supporting bio-efficacy testing) can provide a proxy for epidemiological outcomes of cRCTs.

## STUDY SPECIFIC OBJECTIVES

### Net durability

- To assess the A.I. activity and durability of each of the dual-A.I. LLINs compared to standard LLINs against susceptible and resistant *An. gambiae* colonies, over 3 years of continuous use by households (HHs) under field conditions.
- To determine and compare physical durability (survivorship and fabric integrity) of the dual-A.I. LLINs and standard LLINs over the 3 year period via a prospective cohort study.
  - Obtain detailed information on factors influencing the development of holes in the nets such as handling, washing, and the environment.
  - Assess net user’s perception of net condition and usefulness.
  - Correlate user perception and physical condition to determine physical net status at which a net is considered “useless” or discarded.

- To assess A.I. content in each of the 4 LLINs under evaluation over 3 years of use by HHs.
- To report adverse events that may be associated with each of the LLINs.

### Efficacy and parameterization of models

- To assess entomological efficacy of each of the dual-A.I. LLINs compared to standard LLINs sampled from cRCT households annually over 3 years, in experimental huts against free flying *An*. gambiae sensu lato (s.l.) and *An. funestus* s.l. to generate more realistic entomological outcomes than those achieved using standardized washing.
- To incorporate outcomes generated by the experimental hut trials in a mathematical model of malaria transmission to establish whether these entomological outcomes can predict the epidemiological effect of the cRCT.

## METHODS AND ANALYSIS

### Study area

The WHO Phase III durability study is part of a four-arm cRCT carried out in Misungwi district (2°51’00.0”S, 33°04’60.0”E), on the Southern border of Lake Victoria, Tanzania. The cRCT study area includes 72 villages, 42,314 households and a population of 251,155 based on a census done in 2018 as part of the study. A detailed description has been provided elsewhere.^8^

Six experimental huts have been constructed in Magu district, Mwanza region, Tanzania (2°34.673’S, 33°07.170’E). The hut study site is situated north of the cRCT study area (Figure 1). The main vectors in this area are *Anopheles funestus* sensu stricto (s.s.), *Anopheles arabiensis* and *Anopheles gambiae* s.s.

**Figure 1:**
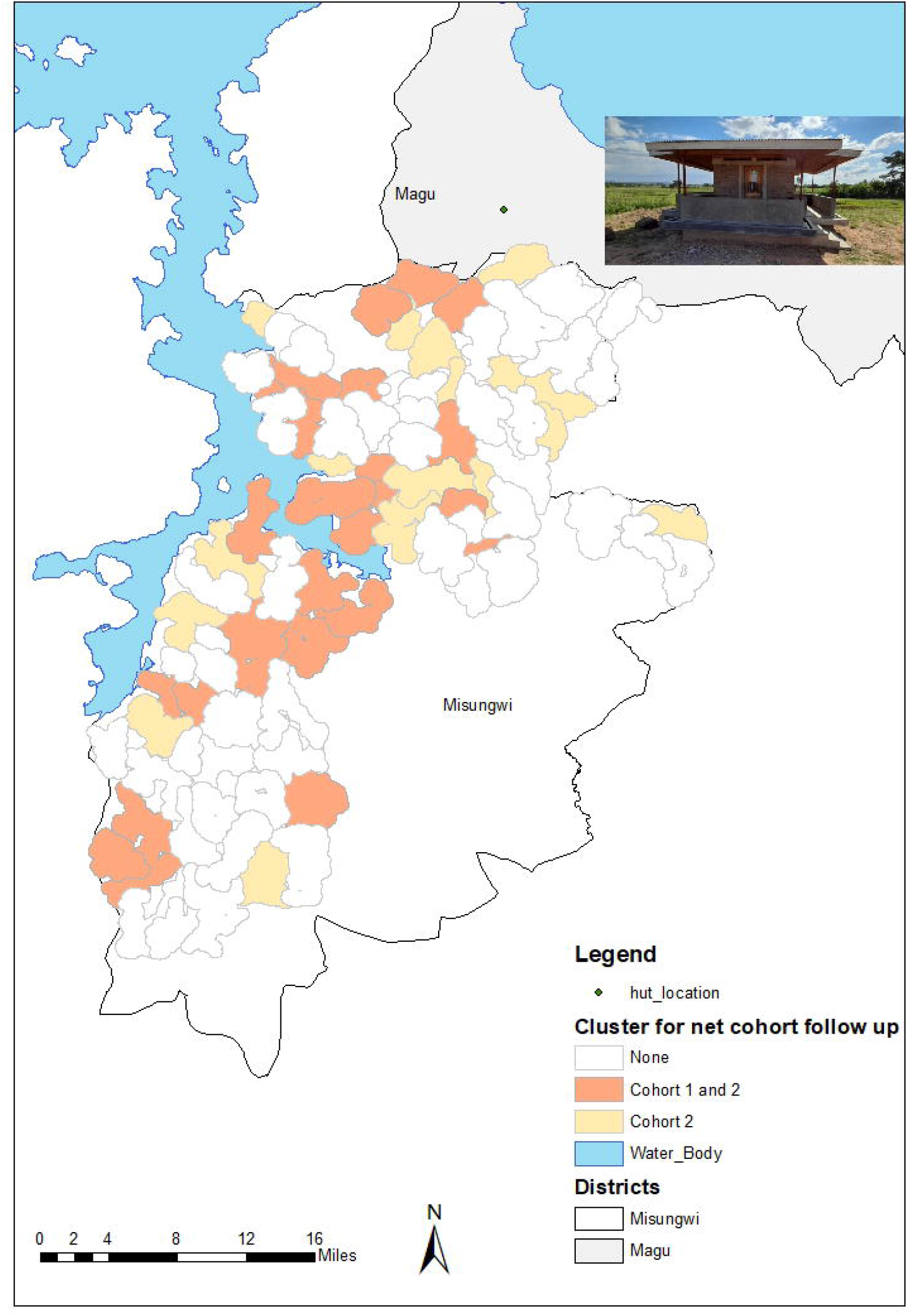
Map showing cRCT site and experimental hut site.

### Intervention

The four LLINs under evaluation are 1/ Royal Guard®, a net combining pyriproxyfen (PPF), which is known to disrupt female reproduction and fertility of eggs, and the pyrethroid alpha-cypermethrin; 2/ Interceptor G2®, a mixture net incorporating two adulticides with differing modes of action; chlorfenapyr and a pyrethroid (alpha-cypermethrin); 3/ Olyset™ Plus an LLIN which incorporates a synergist, piperonyl butoxide (PBO), to enhance the potency of pyrethroid insecticides and; 4/ Interceptor® an alpha-cypermethrin only LLIN and the reference intervention (Table 2).

**Table 2:**
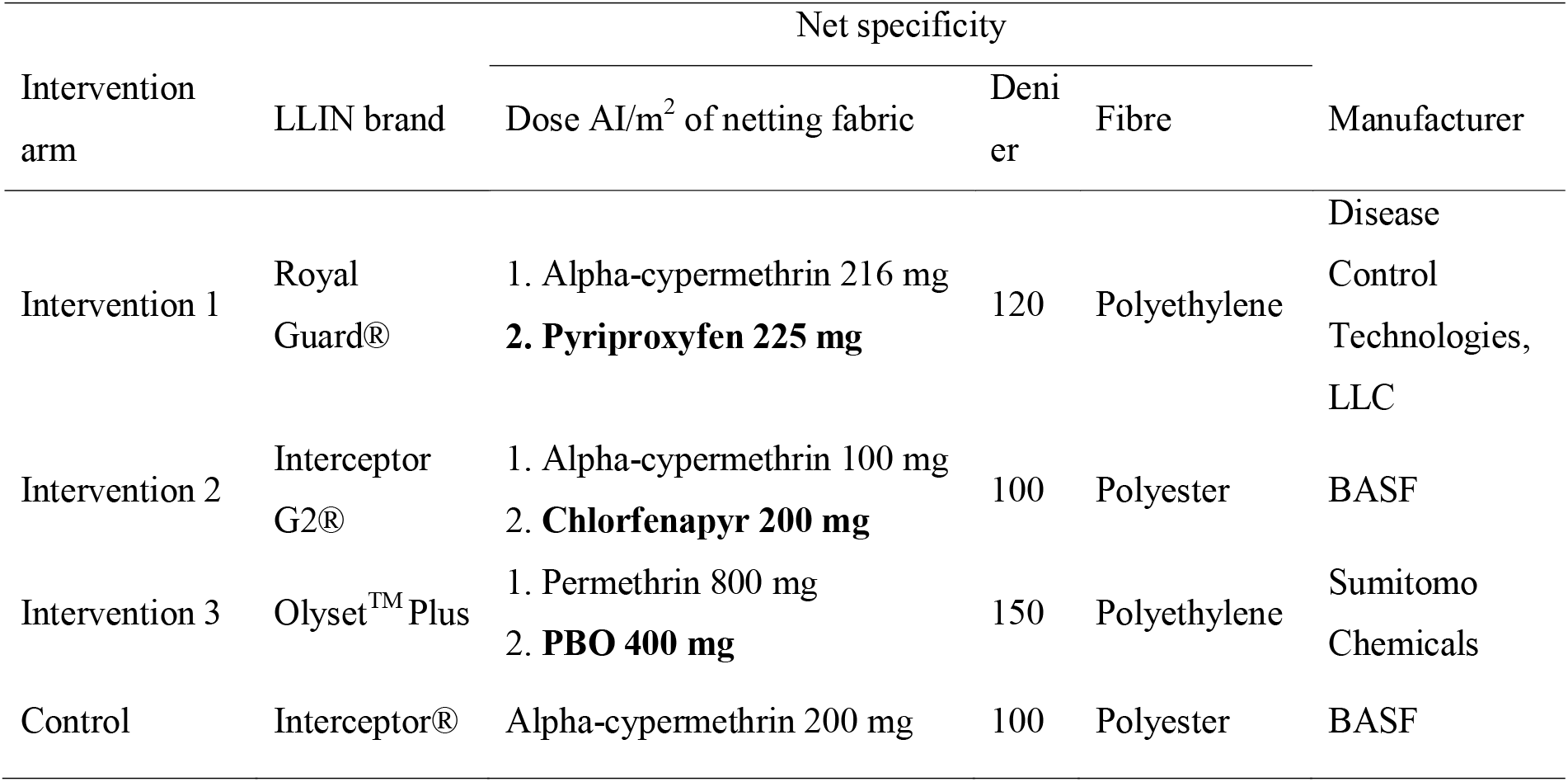
Characteristics of the different type of LLINs under evaluation.

In February 2019, each of the four study LLIN types under evaluation were distributed to 21 clusters (overall number of clusters in the study area which received LLIN were 84) based on restricted randomisation allocation. Each household received one net for every two people.

All nets distributed were blue and rectangular (180 cm length×160 cm wide×180 cm high). In each intervention arm, net survivorship and fabric integrity (cohort 1) are followed in 250 households, selected nearby each other in 5 clusters, and nets for bio-efficacy, chemical analysis and experimental hut trials (cohort 2) sampled from 650 households in 10 clusters (Figure 2).

**Figure 2:**
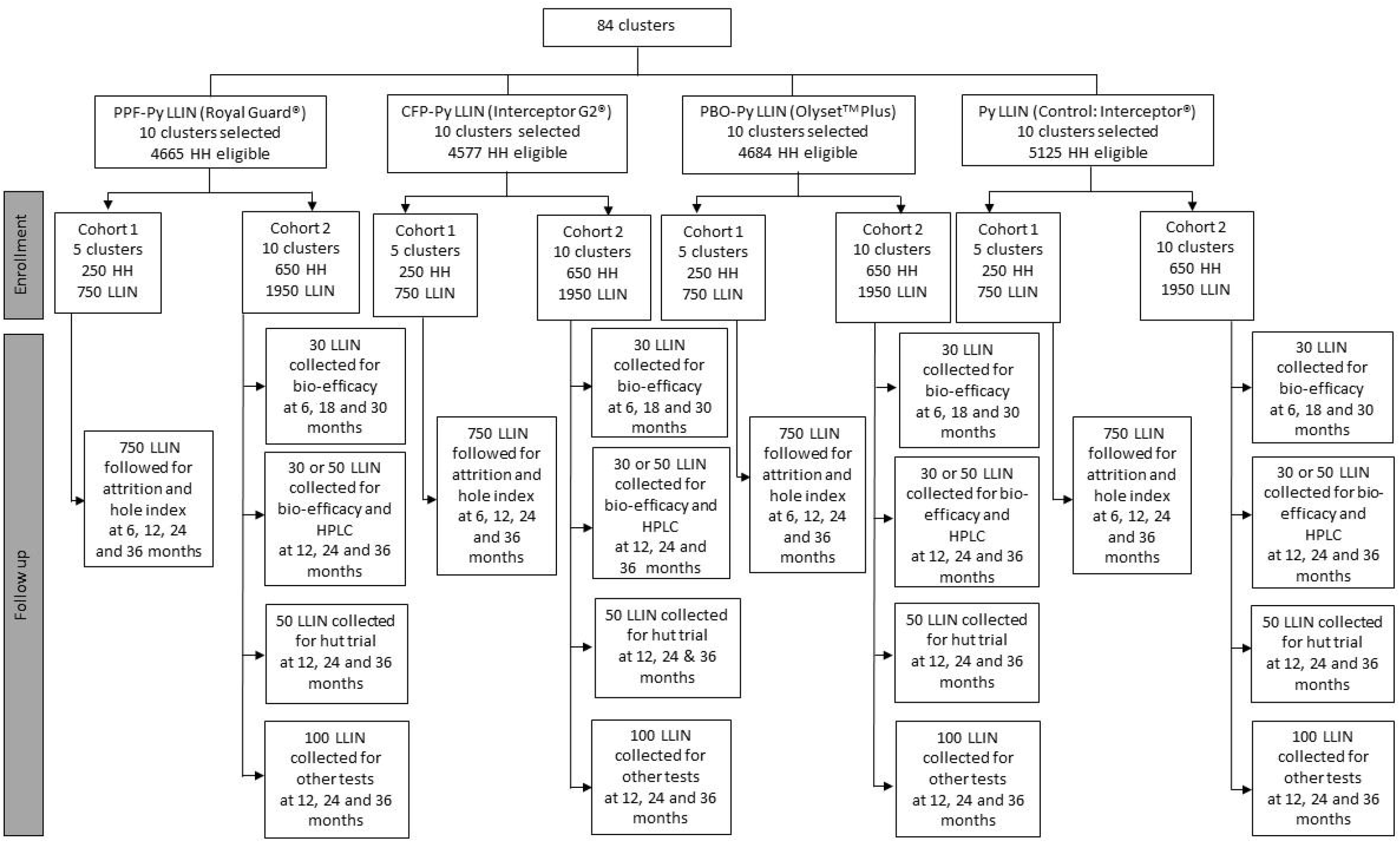
Study design flow chart

### Study Outcomes

Study outcomes are summarized in Table 3.

**Table 3:**
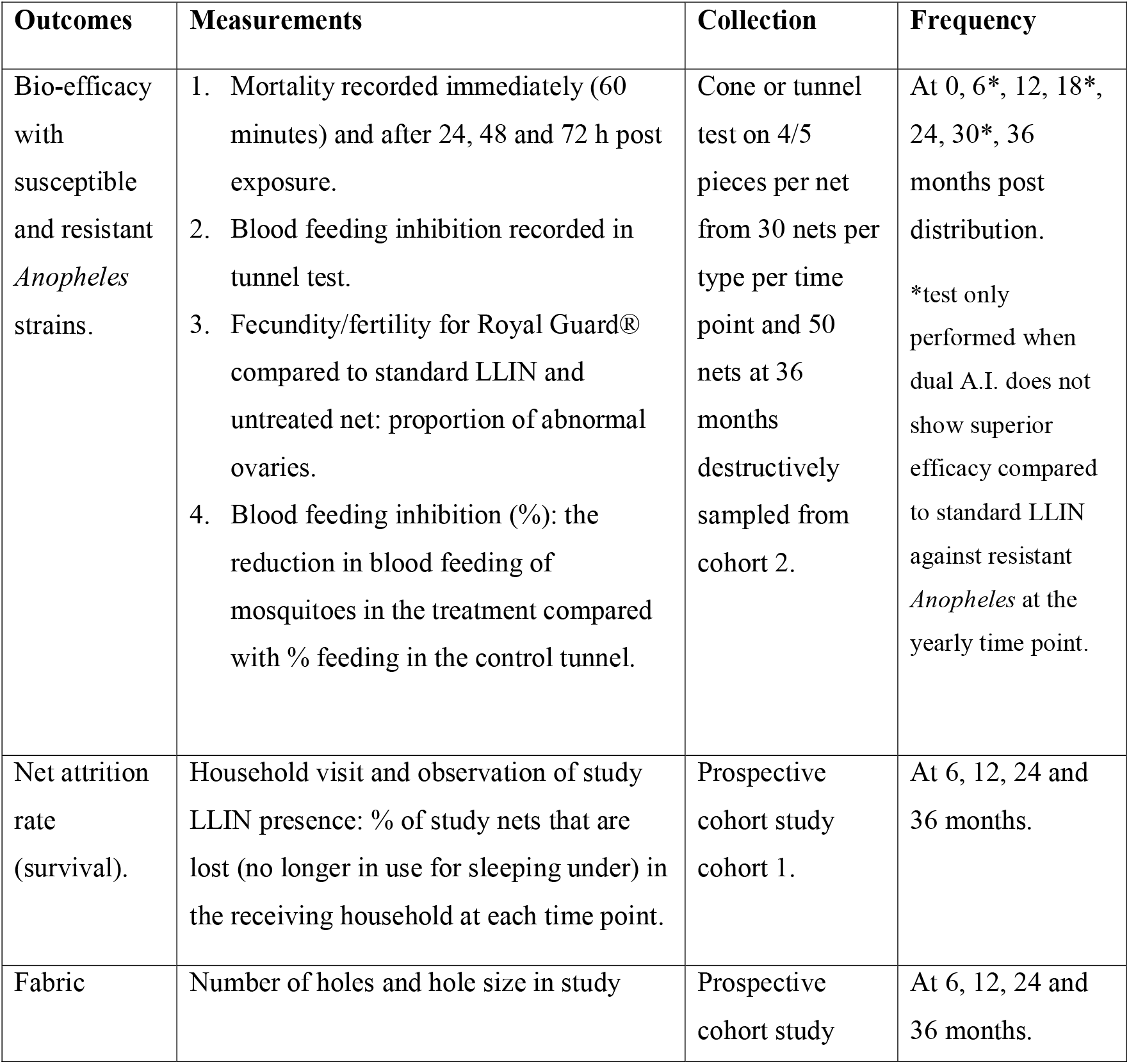
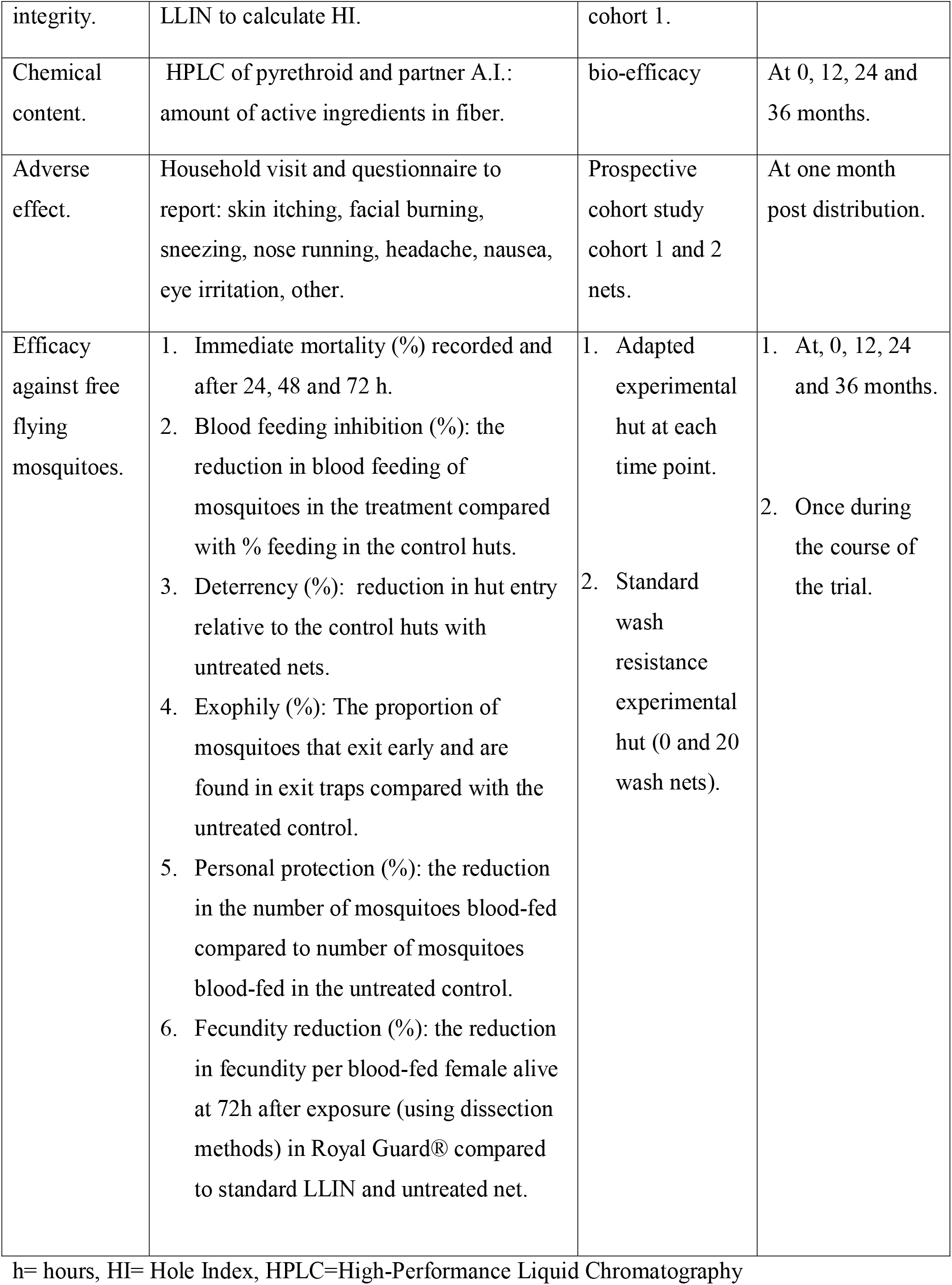
Study outcomes measurements.

### Study design: Net durability

The efficacy and physical durability of the nets will be evaluated by means of a prospective cohort study (Figure 2). A census/enumeration of the household in the hamlet has been complete as part of the cRCT and for each houses name and GPS coordinate are available.

#### Net attrition and fabric integrity (cohort 1)

A total of 250 HHs in 5 clusters per arm will be followed for LLIN physical integrity and attrition. Assuming an average of 3 LLINs per HH based on the average number of sleeping places, this will yield 750 LLINs. All 750 LLINs will be followed for attrition and for hole index. In each cluster HHs will be selected close to each other. During the first visit one month post distribution, in all the households willing to participate used LLIN will be identified with a unique identification number. Household information on quality of housing and other socio-demographic characteristics will be collected (supplementary file 1). The physical presence of all LLINs in the 250 HHs will be recorded at each visit; 6, 12, 24 and 36 months. If the net is still present in the HH, the investigator will record whether the net is being used for its intended purpose. Nets that are not used anymore will also be recorded. If the net is no longer in the house, the investigator will determine how it was lost by asking probing questions. All LLINs followed for attrition and in use will be inspected for, number of holes (including tears in the netting and split seams) by location on the net and size will be classified into four categories: smaller than a thumb (0.5–2 cm), larger than a thumb but smaller than a fist (2–10 cm), larger than a fist but smaller than a head (10–25 cm) and larger than a head (> 25 cm).

Evidence of repairs to the net fabric and the type of repair will be recorded (supplementary file 1). Hole counts will be made by removing each net and arranging it over a frame and returning the nets after measuring physical integrity in the field.

#### Net withdrawal for insecticidal residual activity assessment and experimental hut trials (cohort 2)

LLINs will be withdrawn at different time intervals to be used for bio-efficacy testing and experimental hut trials. To reduce the impact of this withdrawal on the cRCT outcomes, 10 clusters (5 clusters used for attrition and integrity and 5 additional ones) per arm will be selected at random. A total of 650 HHs will be selected at random to give approximately 1950 LLINs per type to follow (Figure 2). The unit of observation will be the individual net.

As for cohort 1 selected nets for cohort 2 will be labelled and household information collected.

From each of the 30 LLINs selected for bio-efficacy, one net piece of 30 cm x 30 cm will be cut from each side at baseline (0 month); in subsequent follow ups only position 2 to 5 will be cut, as position 1 situated at the bottom of the net may be exposed to extreme abrasion when tucked under the bed (Figure 3). At each position, 3 samples adjacent to each other will be removed. The first one will be used for chemical assay (High-Performance Liquid Chromatography; HPLC) and other two for bio-efficacy (one for exposure to *An. gambiae* s.s. strain (Kisumu) and the other for exposure to a resistant strain).

**Figure 3:**
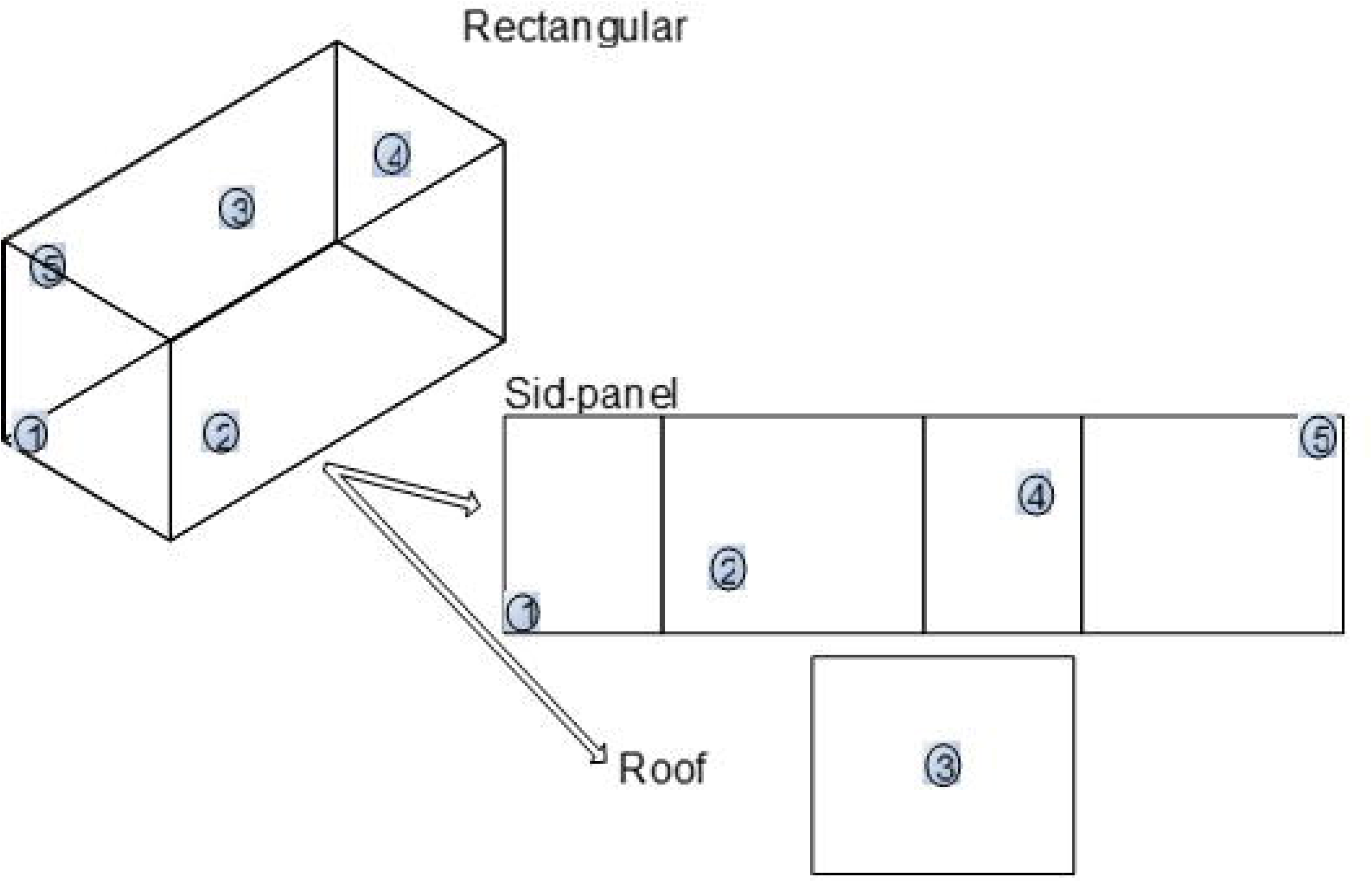
LLIN sampling pattern taken from WHO Guidelines for laboratory and field-testing of long-lasting insecticidal nets.

#### Adverse events

For 200 HHs per intervention (20 HHs per clusters), we will also record perceived adverse effects from users or guardian of users of one study LLIN. The HH will be selected at random from all enrolled in cohort 1 and 2.

### Study design

Net bio-efficacy

#### Bio-efficacy: mosquito strains

In the context of bioassay testing of dual-A.I. LLINs, we will follow the WHO (2013) and (2017) guidelines on modifications to testing, where the evidence suggests this is justified to improve data value or to generate new evidence to inform testing policy.^10 21^

A susceptible *An. gambiae* s.s. strain (Kisumu) will be used to assess the bio-efficacy of the pyrethroid in each of the dual-A.I. LLIN. To assess the durability (bioavailability) of the partner insecticide, it will be necessary to use a pyrethroid resistant strain or species ideally with resistance intensity great enough to withstand the effect of the pyrethroid. We will use the *An. gambiae* s.s. Muleba-kis pyrethroid resistant strain (characterized by both *kdr*-East L1014S and mixed function oxidases-based resistance) to assess the partner A.I. The strain will be kept under constant pyrethroid selection pressure and phenotypic and genotypic resistance will be monitored regularly to assess changes in resistance frequency and intensity. The selection will be done once per each generation at larval stage using 0.08 µg/ml of alpha cypermethrin. Control larval bowls will be treated with 1ml of ethanol.

#### Bio-efficacy: testing methods

##### Rationale

In the dual-A.I. LLIN Interceptor G2®, alpha-cypermethrin is fast-acting, and following a short exposure in contact bioassays, susceptible mosquitoes are knocked down within 1h and killed within 24h. The pyrrole chlorfenapyr requires a longer exposure, is slow-acting and takes up to 72h to kill following a contact bioassay; for delayed mortality, the WHO has proposed that mosquitoes may be held up to 72h with mortality reported every 24h. There is clear evidence that cone bioassays with 3 min exposures fail to predict field efficacy of Interceptor G2®.^22^ WHO has stated that tunnel tests may be more appropriate to estimate the field durability of chlorfenapyr.^23^

In the dual-A.I. LLIN Royal Guard®, PPF impacts egg development, while alpha-cypermethrin will induce mortality. The reproductive effects of PPF on blood-fed female mosquitoes are 3-fold. The first effect of PPF exposure is to disrupt the maturation of eggs and oviposition by females 2-3 days after blood-feeding. The second observable effect is reduction in the mean number of eggs per ovipositing female. The third effect is reduction in the hatch rate of laid eggs or production of viable larvae. Conventionally, the effects on reproductive outcomes are assessed by observation of oviposition rate and hatch rate in mosquitoes exposed to PPF compared to unexposed mosquitoes.^24 25^ The problem with oviposition as an indicator of fertility/sterility is the low oviposition rate in control, unexposed mosquitoes. Furthermore, direct observations require a long follow-up, appropriate infrastructure and can be laborious. An alternative approach is to dissect mosquitoes after exposure when eggs should normally have become fully mature (2-3 days post-blood meal). During the normal gonotrophic cycle, after taking a blood meal, the mosquito’s oocytes change in size and shape, and finally reach Christopher’s stage V which are a distinctive crescent shape (Supplementary file 2).^26^ Previous work on PPF-treated females has shown morphological defects on oocyte maturation in the ovaries following exposure, with development of PPF-affected oocytes arrested before reaching stage V.^27^

In the dual-A.I. LLIN Olyset™ Plus, PBO inhibits the cytochrome P450 oxidases responsible for metabolic pyrethroid resistance while permethrin will induce mortality. Cone tests have been used effectively to assess the performance of Olyset™ Plus^28^ however, there is a need to use a resistant strain with oxidase-based mechanisms for these bioassays to assess field durability of PBO.

##### Cone test

WHO cone tests will be performed on each of the pieces of the 30 (t0-t30) and 50 LLINs (t36) collected for the following product Royal Guard® and Olyset ™ Plus (both with susceptible and resistant mosquito strain) and Interceptor G2® (only against susceptible strain).^10^ For each net, all pieces cut will be tested. Four replicates will be done per net piece using 5 mosquitoes per cone for a total of 20 mosquitoes per net piece and 80 mosquitoes per net (100 for the baseline). An untreated net will be run in parallel during each testing as well as 2 pieces of the standard LLIN collected at the same time point. The estimated number of mosquitoes to be tested per time point is detailed in Table 4.

**Table 4:** Summary of the bioassay testing plan and outcomes per net type and mosquito strain.

|  |  | **Interceptor G2®** | **Royal Guard®** | **Olyset™ Plus,** |
| --- | --- | --- | --- | --- |
| **Susceptible strain: Assessment of pyrethroid** |  |  |  |  |
|  | Treatment | 1/Untreated,<br>2/Interceptor,<br>3/Interceptor G2® | 1/Untreated,<br>2/Interceptor,<br>3/Royal Guard® | 1/Untreated,<br>2/Interceptor,<br>3/Olyset™ Plus |
| Cone | Outcomes | Kd*, mortality 24, 48<br>and 72 h** | Kd, mortality 24, 48<br>and 72 h | Kd, mortality 24, 48 and 72<br>h |
|  | Exposure time | 3 minutes | 3 minutes | 3 minutes |
|  | Total nets | 30 nets (t0 to t30), 50<br>nets (t36) | 30 nets (t0 to t30), 50<br>nets (t36) | 30 nets (t0 to t30), 50 nets<br>(t36) |
|  | Total pieces | 5 (t0) and 4 subsequent<br>follow ups (position 2<br>to 5) | 5 (t0) and 4 subsequent<br>follow ups (position 2<br>to 5) | 5 (t0) and 4 subsequent<br>follow ups (position 2 to 5) |
|  | Total replicates /<br>piece | 4 | 4 | 4 |
|  | Total Mosquito<br>per test | 5 | 5 | 5 |
|  | Total<br>mosquitoes | Total between 4800 to<br>6000 per time point | Total between 4800 to<br>6000 per time point | Total between 4800 to 6000<br>per time point |
| Tunnel | Outcomes | Kd, mortality 24, 48<br>and 72 h, blood feeding | Kd, mortality 24, 48<br>and 72 h, blood<br>feeding | Kd, mortality 24, 48 and 72<br>h, blood feeding |
|  | Exposure time | 12 to 15 h | 12 to 15 h | 12 to 15 h |
|  | Total nets | All failed net with cone | All failed net with cone | All failed net with cone |
|  | Total pieces | 1 per net | 1 per net | 1 per net |
|  | Total replicates /<br>piece | 2 replicates | 2 replicates | 2 replicates |
|  | Total Mosquito<br>per test | 50 | 50 | 50 |
|  | Total<br>mosquitoes | Determined by number<br>of failing nets | Determined by number<br>of failing nets | Determined by number of<br>failing nets |
| **Resistant strain: Assessment of partner A.I. or synergist** |  |  |  |  |
| Cone | Outcomes |  | Kd, mortality 24, 48 and 72 h, ovarian development | Kd, mortality 24, 48 and 72 h |
|  | Exposure time |  | 3 minutes | 3 minutes |
|  | Total nets |  | 30 nets (t0 to t30), 50 nets (t36) | 30 nets (t0 to t30), 50 nets (t36) |
|  | Total pieces |  | 5 (t0) and 4 (t6 to t36) | 5 (t0) and 4 (t6 to t36) |
|  | Total replicates / piece |  | 4 | 4 |
|  | Total Mosquito per test |  | 5 | 5 |
|  | Total mosquitoes |  | Total between 4800 to 6000 per time point | Total between 4800 to 6000 per time point |
| Tunnel | Outcomes | Kd, mortality 24, 48 and 72 h, blood feeding | Kd, mortality 24, 48 and 72 h, Blood feeding, ovarian development | Kd, mortality 24, 48 and 72 h, blood feeding |
|  | Exposure time | 12 to 15 h | 12 to 15 h | 12 to 15 h |
|  | Total nets | 30 nets (t0 to t30), 50 nets (t36) | All failed net with cone | All failed net with cone |
|  | Total pieces | 1 per net (position 2) | 1 per net | 1 per net |
|  | Total replicates / piece | 2 replicates | 2 replicates | 2 replicates |
|  | Total Mosquito per test | 50 | 50 | 50 |
|  | Total mosquitoes | 30 (50) nets x 1-piece x 2 replicates x 50 mosquitoes x 3 treatments = 9000 | Determined by number of failing nets | Determined by number of failing nets |
|  | Net Specificity | slow killing effect 72 hours; mortality main outcome | Ovarial development by dissection at 72 h post exposure | Colony mosquito's resistance strain (kdr*** and kdr+MFO****) |

\* knockdown, \*\* hours, \*\*\*knockdown resistance, \*\*\*\* cytochrome P450 mono-oxygenase mechanisms

Per bioassay, five unfed 2-5 days old *An. gambiae* s.s. will be introduced into each cone. After 3 min exposure, mosquitoes will be transferred into labelled paper cups covered with untreated netting with access to 10% sugar solution. The bioassays will be carried out at 25±2°C and 75±10% RH and at 27±2°C.

##### Tunnel test

LLIN pieces (Interceptor G2®, Royal Guard® and Olyset ™ Plus,) that do not meet the criteria of ≥95% knockdown (kd) rate after 60 minutes and/or a mortality of ≥80% after 24 hours in cone bioassays (all pieces and replicates for the same net pooled together), will be evaluated in a tunnel test to determine the efficacy (mortality and blood feeding inhibition) using a guinea pig as the bait. One netting piece for each failing net will be tested in the tunnel in two replicates. The net piece that produces mortality closest to the mean mortality during the cone test will be used in the tunnel test. Because the cone is not suitable for initial testing of Interceptor G2® using the resistant strain, the piece in position 2 of each of the 30 nets will be systematically tested in the tunnel (Table 4).

The procedure for use of guinea pigs will be compliant with criteria laid down in EC Directive 86/609/ECC concerning protection of animals used for experimental purposes. The animal ethics approval has been sought from LSHTM. The glass tunnel is 25 cm^2^ and 60 cm long, divided at one third of the length by a disposable cardboard frame to which the LLIN netting piece is attached. The surface of netting “available” to mosquitoes is 400 cm^2^ (20 cm x 20 cm). Nine holes, each 1 cm in diameter, (one at the center of the square and the other eight equidistant at 5 cm from the border) will be made in the netting to allow for passage of mosquitoes. Netting-covered cages at both ends provide easy access to add and remove mosquitoes. In one cage, a guinea pig will be restrained. Fifty mosquitoes will be introduced at the opposite end of the tunnel from where the guinea pig is restrained. The experiment will begin at 18:00 and end at 08:00 the following morning, mosquitoes will be scored according to whether they passed through the netting, whether they successfully blood fed and whether they survived the exposure period.

##### Cylinder test

The cone and tunnel tests may prove inadequate for evaluating the durability of partner A.I.s over 1-3 years once the insecticidal content starts to decrease. The standard cone test exposes mosquitoes for 3 min only, which has been shown to underestimate contact time and the exposure time and mortality attained in experimental huts using resistant mosquitoes; 30 min exposure is probably more realistic when using resistant mosquitoes.^29^ Mortality generated for free flying resistant mosquitoes in experimental huts correlates well with mortality attained in 30 min bioassay for some insecticides tested, e.g., chlorfenapyr and Interceptor G2® (Rowland and Kirby unpublished data). While the contact time may differ for other insecticides or when the concentration decreases during 3 years in the field; now that the precedent is established for one type of dual-A.I. LLIN, the average contact time of free flying mosquitoes is likely to be longer than 3 min for other nets too. The WHO cylinder test will be performed for Olyset™ Plus, Royal Guard® and Interceptor G2® on a sub-sample of nets at each time point and compared to tunnel and cone results.

The netting will be stapled to WHO control test papers measuring 15cm x 12cm to facilitate rolling and fitting into the test cylinder in the same way as an insecticide test paper would be fitted. Holding rings are inserted to hold back the netting.^30^ Bioassays will follow the same procedure as insecticide susceptibility testing except that exposure time will be 3, 15, 30 and 60 min as necessary. Before exposure, 10 mosquitoes will be aspirated into the holding cylinder of the kit and then blown into the exposure cylinder according to standard procedures. After exposure, the test insects are blown back into the holding cylinder and 10% sugar solution provided. Ten mosquitoes per cylinder test would ensure a density per unit area of netting similar to that of five mosquitoes per cone.

### Experimental hut design

The experimental huts are a modified version of the standard East African hut,^31^ featuring four brick walls, a wooden ceiling lined with hessian sackcloth, an iron sheeting roof, two baffled eave gaps above each wall, and a window trap on each wall. The huts are built on concrete plinths and surrounded by a water-filled moat to deter entry of scavenging ants. In the modified design, the four verandas are open; the baffled eave gaps above all four sides allow unimpeded entry of mosquitoes and minimal mosquito exiting. Mosquitoes are restricted to exiting through the window traps on the four walls of the hut.

In each hut, cloth sheets are laid on the floor each night to ease the collection of knocked-down mosquitoes in the morning. Sugar solution is provided at night in the window traps to reduce mosquito mortality.

The nets will be evaluated using experimental huts for their effects on free-flying, wild *An. gambiae* s.l., *An. funestus* s.l. and *Culex quinquefaciatus* mosquitoes for their ability to deter entry, repel mosquitoes, induce mortality and inhibit blood-feeding.

#### Adapted experimental hut study

Each of the 30 individual nets per product type collected from community at t12, 24 and 36 months will be tested in experimental hut. The following treatment shall be assessed at each time point:

1. Control: untreated polyethylene net with 6 holes
2. Standard LLIN: new interceptor washed one time with 6 holes
3. Interceptor at t12/t24/t36
4. Interceptor G2® at t12/t24/t36
5. Royal Guard® at t12/t24/t36
6. Olyset™ Plus at t12/t24/t36

The study will be done over a 6 week periods. Sleepers will be rotated between huts on successive nights to account for individual attractiveness and net treatment every week following a random Latin square design (supplementary file 3). Every week, collections shall be performed over 6 days and the last day will be used for cleaning and airing before the treatment rotation. Six replicates of untreated net and new standard LLIN will be tested per hut treatment and will be swapped every day within each week of the trial. Field collected nets will be changed every day and tested for one night only per trial. Because there are 36 day/night collection per treatment for a complete Latin square rotation (sleepers and treatments) and only 30 field collected individual nets per product type an additional 6 new LLIN from each treatment will be evaluated (treatment 3 to 6). Six holes of 4 × 4 cm will be cut in the untreated and new LLINs used in each treatment arm following WHO guidelines.^10^ Hole size will be counted for the field collected nets as for cohort 1 nets followed for fabric integrity. A hut trial study for each time point will be repeated 2 to 4 times to account for vector composition seasonality.

#### Mosquito processing

All mosquitoes collected in experimental huts will be monitored for three days and mortality recorded after 24, 48 and 72h post collection. Blood fed mosquitoes collected from Royal Guard®, Interceptor® and untreated nets, will be dissected after 72h for observation of fertility/sterility. Mosquitoes will be recorded as fertile if the eggs are fully developed into Christopher’s stage five.

A subset of alive and dead mosquitoes from each hut will immediately be killed (if still alive) and stored in RNAlater® at -80°C for species identification and resistance gene expression analysis. Following molecular species identification, presence/absence of resistance alleles will be compared between individuals of assumed resistant (alive) and susceptible (dead) phenotypes and changes in allele frequency will be compared between hut conditions and between baseline characterization and post-intervention.

#### Modeling of experimental hut trial entomological surrogates

While experimental hut trials are the gold standard for assessing LLIN efficacy against mosquitoes, mathematical modelling^2 32^ can be used to predict the public health impact of factors such as pyrethroid resistance on LLIN efficacy and malaria transmission.^2^ The models are calibrated to the local area using site specific entomological and epidemiological data collected from the cRCT site (such as baseline mosquito bionomics, history of LLIN use and baseline malaria prevalence). Two sets of parameters are used to characterise the efficacy of trial dual-A.I. LLINs. The first uses estimates of the proportions of mosquitoes dying, blood-feeding and outcomes such as deterrence, exiting and repellence estimated using experimental hut trials conducted in the region of the cRCT. The second uses estimates for the same metrics derived from a meta-analysis of all currently available experimental hut trial data for the same dual-A.I. LLINs from across Africa. Models parameterised with these two sources of data are used to predict changes in malaria prevalence over time. These two models are statistically compared to the observed results of the cRCT at different time points following the mass campaign to investigate the benefit of local LLIN efficacy information. WHO discriminating dose bioassays are used to quantify the frequency of resistance in the mosquito populations in the vicinity of the experimental hut site to provide a link between the outcomes of the trial and widely used assays for assessing the frequency of resistance.

There is considerable uncertainty in how the efficacy of nets changes over time. It can be estimated using the WHO proxy of standardised washing, as described by experimental hut outcomes from trials evaluating LLINs washed 0 and 20 times. The number of washes taken to halve the killing activity of the LLIN can be estimated and converted into predictions of the insecticidal half-life in years considering 20 washes to represent the decay expected in an LLIN over three years of use in the field. Estimates of the actual duration of insecticidal activity in the field is one of the more uncertain features of experimental hut trials and consequently for models evaluating the public health impact of novel LLINs. This is because of the uncertainty in the relationship between the number of washes and durability of insecticide in the field over time for non-pyrethroid A.I.s. This uncertainty is important to include as the average age of LLINs in Africa is over one year old and small changes in LLIN-induced mortality over time can have a large epidemiological impact as population LLIN coverage falls two or three years after the last mass distribution campaign. This problem seems particularly acute for Interceptor G2® as chlorfenapyr cannot be evaluated in simple cone bioassays and nets washed 20 times.^33^ Moreover, LLINs used in experimental hut trials sometimes appear indistinguishable from unwashed nets whereas under field condition nets are subject to many environmental factors affecting durability in addition to washing, such as friction, wood smoke, and everyday wear and tear. Evaluation of naturally aged LLINs collected from the field in experimental hut trials may allow more precise predictions of the longevity of a LLIN (i.e. half-life), allowing the decay in insecticidal activity to be directly estimated instead of having to rely on proxy measures. This is anticipated to improve the accuracy of epidemiological predictions made from entomological data which can be evaluated by comparing model predictions to the results of the main cRCT. The utility of incorporating other entomological data collected as part of the trial into the modelling framework shall be investigated.

## SAMPLE SIZE CALCULATION

Sample size calculations for prospective LLIN study of net survivorship were performed using the power log rank command in Stata v.15.1. A total of 750 LLINs per type from 5 clusters per arm (i.e. 150 per cluster) will allow detection of a 9.4% absolute difference (hazard ratio = 0.8651) in LLIN attrition rate if we assume an attrition rate in the control arm of 70% over the 3 years. This is assuming an intra-cluster correlation coefficient (ICC) of 0.03. We can detect a hazard ratio=0.7951 (i.e. 14.3% absolute difference) if we assume an ICC=0.01, and a hazard ratio=0.7478 (i.e. 17.7% absolute difference) for an ICC=0.02.

## DATA MANAGEMENT & ANALYSIS

### Data management

All data on LLIN physical conditions, washing and household characteristics will be collected using Open Data Kit (ODK) forms. Bioassay data will be recorded on standardised forms and double entered into an Access file and linked to the database via the net identification number and time interval. Consistency checking will be done by running algorithms especially designed to identify sources of error. This database will be sent by the data manager to the project manager after each time-interval. The project manager will keep an updated master list of the location and status of each net. Data from bioassay and chemical residue analysis will be entered separately.

Hut data will be entered directly in electronic forms prepared in ODK. Standard Operating Procedures (SOPs) for data collection will be developed and field staff will be appropriately trained to ensure rigorous data collection. This will include quality control (QC) of their own performance by checking for missing data or implausible responses. Further QC will be conducted by a supervisor who monitors performance of field staff by checking for completeness and internal consistency of responses within hours of data collection. To maintain participant confidentiality, all consents forms will be kept in a locked cabinet only accessible by an authorized staff. Statistical analysis will be performed using Stata software v.15.1.

### Data analysis

#### Fabric integrity (hole index)

Each of the hole categories will be weighted according to WHO guidelines.^10^ HI will be calculated using the formula HI□=□(1□×□no. of size-1 holes)□+□(23□×□no. of size-2 holes)□+□(196□×□no. of size-3 holes)□+□(576□×□no. size-4 holes). Based on the HI the nets will be divided into 3 categories 1/ Good: HI <64, 2/ Acceptable: pHI between >64□<□=642, 3/ too torn; pHI >642. Good and acceptable nets will be combined into a serviceable condition category.

#### Attrition/survival

The rate of attrition will be first calculated as the proportion of study nets lost among all study nets originally received and further divided into reasons of net loss. To estimate functional survival only the attrition due to destruction, discarding or use for other purpose will be included (MPAC recommendation).^34^ The functional survival rate (pX) of each type of study net at each time point will be calculated as:

**pX**=% surviving to time X **=** (number of study net present and “serviceable” at time X **/** number of study net originally received and not given away at time X) * 100 Descriptive statistics will be used to present the proportion surviving for each study net at each time point and compare the dual-A.I. LLINs to the standard LLIN. Cluster effect will be accounted for to estimate the 95% confidence interval.

#### Median survival time

Median survival time is the time point at which 50% of the study nets received are still present and in serviceable condition. First the functional survival rate to time X will be compared against a reference survival curve provided by WHO in the VCTEG report.^34^ The functional survival rate to time X will also be compared between the dual-A.I. LLINs against the standard LLIN. A Kaplan–Meier survival analysis will be used to estimate the median survival time of each study net and will consider the design effect.

#### Efficacy and long-lasting effect of the pyrethroid

The dual-A.I. net will meet WHO criteria^10^ for efficacy and long-lasting effect of the pyrethroid if, after 36 months of use, at least 80% of sampled nets tested against a pyrethroid susceptible mosquito strain are effective in WHO cone tests (≥ 95% knockdown or ≥ 80% mortality after 24 hours) or tunnel tests (≥ 80% mortality or ≥ 90% inhibition of blood-feeding). Thresholds for the second A.I. of each dual-A.I. LLIN are not known yet and would need to be established in relation to the cRCT findings. However, dual A.I. LLIN efficacy outcomes against resistant *Anopheles* strain will be compared to those of standard LLIN to assess superior effect at each time point.

#### Experimental hut trial analysis

Proportional outcomes (blood-feeding, exiting and mortality, oviposition) related to each experimental hut treatment will be assessed using binomial generalized linear mixed models (GLMMs) with a logit link function. A separate model will be fitted for each outcome. In addition to the fixed effect of each treatment, each model will include random effects to account for the following sources of variation: between the huts; between the sleepers; between the weeks of the trial; between trial rounds; and finally, an observation-level random effect to account for variation not explained by the other terms in the model (over dispersion).

Differences in deterrence, proportions of adult females killed, proportions blood-fed, overall killing effect, personal protection and number of females laying eggs between the treatments will be analysed using negative binomial regression based on numbers entering, numbers blood-fed, killed and numbers observed for oviposition, respectively, with adjustment for the above-mentioned covariates.

Entomological data from the entomological surrogate studies (experimental hut trials and supporting efficacy bioassays) will be integrated into a meta-analysis and modelling of disease outcomes to investigate possible relationships between epidemiological and entomological data from the cRCT.

Comparisons of the changes in levels of gene expression will be performed between live mosquitoes collected from experimental huts containing treated interventions and experimental huts with no treatment/control treatment (relative to a susceptible laboratory control colony). The assumptions are that comparing between these two experimental groups will allow us to differentiate between metabolic genes which are constitutively over-expressed in our field populations and may contribute to resistance (i.e. expression levels observed in the control hut which contains a mixture of ‘resistant’ and ‘susceptible’ vectors of unknown phenotype) and those genes which are over-expressed in our field populations in response to intervention/insecticide exposure (i.e. expression levels observed in the treated huts compared to the control huts).

#### Frequency of Analyses

Analysis of entomological outcomes from the hut trials will be done at the end of each hut trial and the data shared with the modellers to generate predicted outcomes ahead of the cRCT analysis.

## Data Availability

Not applicable.

## ETHICS APPROVAL

This protocol has received ethical approval from the Medical Research Coordinating Committee (MRCC) of the National Institute for Medical Research (NIMR), Kilimanjaro Christian Medical University College (KCMUCO), and the London School of Hygiene and Tropical Medicine (LSHTM).

For the community phase III durability activity written informed consent will be obtained from an adult in the household (Supplementary file 4). Written consent forms will also be obtained from volunteers (sleepers) who agree to participate in the hut study (supplementary file 5). Only adults of 18 years or older will be recruited, excluding pregnant women. Volunteers will be offered daily chemoprophylaxis and the risks of malaria explained (willingness to take will be an inclusion criterion). By sleeping under a mosquito net, they will obtain protection not dissimilar to exposure they would normally obtain against mosquitoes in their own home should they use a pyrethroid LLIN. Because an untreated net is used in one arm, all volunteers will be monitored each day for signs of fever. Confirmed falciparum parasitaemia will be treated with Coartem (artemether 20 mg/lumefantrine 120 mg) by a local physician. No side effects are expected from the LLIN except possibly some irritation to mucous membranes; these will be monitored and reported.

Guinea pigs will be used in the partner Institutions in Tanzania to feed the colony mosquitoes. Restrained guinea pigs are also used in WHO tunnel test as bait. KCMUCo and NIMR laboratories in Tanzania have obtained approval from the Animal Welfare and Ethical Review Board of LSHTM for the use of animals for this study.

## DISSEMINATION

Study findings will be shared in national and international stakeholder meetings. Results will also be shared through presentation to the relevant WHO programmes and through peer-reviewed publications, at scientific conferences and will be reported through clinicaltials.gov.

## COMPETING INTEREST

The authors declare that they have no competing interests.

## AUTHORS CONTRIBUTIONS

JLM contributed to selection of the study area, implemented and supervised the experimental hut construction and drafted the manuscript; LAM contributed to selection of the study area and was a major contributor to the revision of the manuscript; FWM contributed to study design and supported the field activities and revised the manuscript; JFM & EL & AM contributed to the overall management of the project, supported field activities, developed household and net questionnaires for the phase III and revised the manuscript; MK contributed to study design, provided sample size calculation and statistics support; TSC & ESS contributed to the design of the experimental hut study, conception of the model and drafting of the manuscript; NP is the trial principal investigator, designed the study, wrote the study protocol, supported implementation of field activities and revised the manuscript; MR designed the study, wrote the protocol, contributed to selection of the study area, and revised the manuscript. All authors read and approved the final manuscript.

## AVAILABILITY OF DATA AND MATERIALS

Not applicable.

## CONSENT FOR PUBLICATION

Not applicable.

## ACKNOWLEDGEMENT

Special thanks to the community for allowing us to construct our experimental hut in their area, to the project technicians for collection, processing, and identification of mosquitoes, and to the volunteers for agreeing to participate in the study. We would like to thank Jackie Cook and David Macleod to have prepared the random latin square stata do file.

## Notes

### Competing Interest Statement

The authors have declared no competing interest.

### Clinical Trial

NCT03554616

### Clinical Protocols

https://bmjopen.bmj.com/content/11/3/e046664

### Funding Statement

This project is funded by the Bill and Melinda Gates Foundation through the Innovative Vector Control Consortium and under the global health trial, a jointly funded initiative from The Department of Health and Social Care, the Department for International Development, the Medical Research Council and the Wellcome Trust (Grant Ref: MR/R006040/1).

