## Supplementary file 2 for "Durability of three types of dual active ingredient long-lasting insecticidal net compared to a pyrethroid-only LLIN in Tanzania: protocol for a prospective cohort study nested in a cluster randomized controlled trial"

| Misungwi Malaria Trial: Efficacy of different types of bi-treated long lasting insecticidal nets and deployment strategy for control of malaria transmitted by pyrethroid resistant vectors.  **Standard Operation Procedure (SOP) for dissection and estimation of oviposition inhibition in resistance *Anopheles* colony strain after exposure to Piryproxyfen/pyrethroid treated nets** | |
| --- | --- |
| 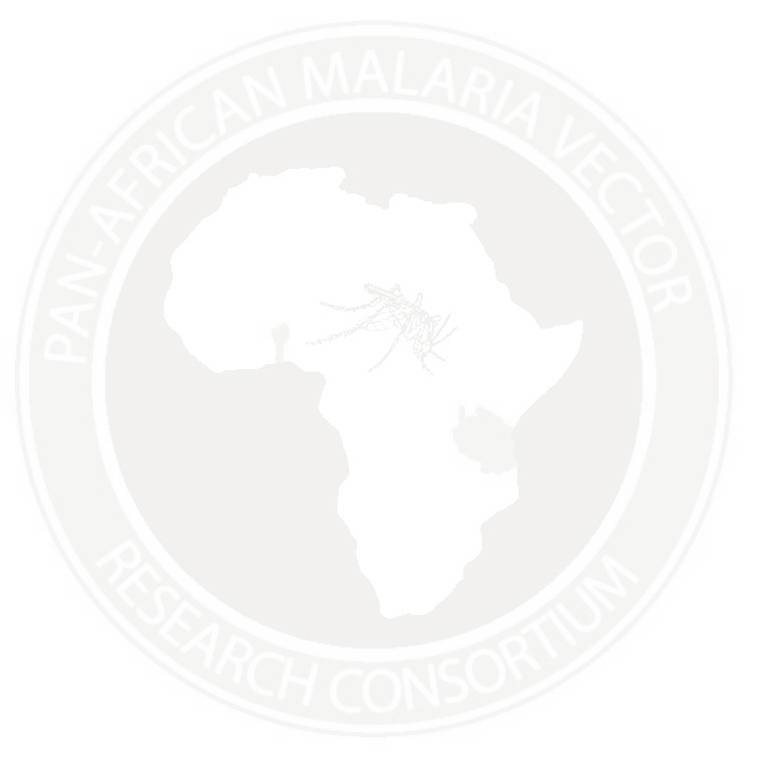SOP Code: | 008 |
| Revision number: | 02 |
| First edition prepared by: | Jackline Martin and Nancy Matowo based on Corey Leclerc and Alina Soto Carson Msc work |
| First edition approved by: | Natacha Protopopoff |
| Revised by: | Joanna Furnival-Adams |
| Date first released: | 06/07/2019 |
| Date last revised: | 21/07/2020 |
| SOP effectives from: |  |
| SOP expires on: |  |
| For use by: |  |
| For use in (Distribute to): | KCMUCO Mabogini insectary testing facility |
| Related forms: | Mosquito dissection form |
| Related documents: | 2014 Methods in Anopheles Research (MR4) |

### 1  Aim

Assess the efficacy of pyriproxyfen incorporated in dual insecticide treated nets on the fertility of resistant Anopheles strain.

### 2 Anopheles tested

Adult female mosquitoes of the *An.gambiae* s.s. Muleba kis (kdr east & MFO) will be obtained from a laboratory colony maintained at KCMUCo. Two to five-day old blood-fed female mosquitoes will be used in bioassays. Wild blood fed resistant mosquitoes of unknown age could also be tested.

### 3 Treatment

- LN Royal Guard
- Untreated Net
- Standard LN: Interceptor

### 4 Outcome measure

- Percentage of dissected females with under-developed ovaries at 72 hours post feeding (i.e. the follicles failed to completely develop from previtellogenic resting stage I to maturity stage V).
- Proportion of dissected females with deformed eggs (eggs not fully developed, at stage II-IV)
- Average number of eggs in the ovaries at 72 hours’ posts feeding.

### 5 Materials for dissection

First prepare all the material for dissection

- Dissecting microscope
- Slides
- Marker pen
- Dissecting kit
- Distilled water
- Compound microscopy
- Lamp
- Record form
- Pen and pencil
- Slide box
- Chloroform
- Cotton wool
- Plastic pipette
- Beaker/paper cup

### 6 Methods

#### 6.1 Cone bioassay

Cone bioassay will be performed according standard WHO procedure. Five freshly blood fed *Anopheles* RSP will be introduced into each cone. Twenty to twenty-five replicates of five mosquitoes will be tested. After a 3 minutes exposure, mosquitoes will be transferred into labelled paper cups covered with untreated netting. When the testing is finished mosquitoes will be provided with 10% sugar solution. Knock down after 60 minutes and mortality at 24, 48, 72 hours will be recorded.

All alive gravid female Anopheles mosquitoes at 72hrs post-exposure will be anesthetized at -20°C freezer condition for five up to ten minutes before dissection. If wild mosquitoes were exposed, prior to dissection, female *Anopheles* species will be morphologically identified by taxa i.e. separating *An. gambiae* sl from *An. funestus* group as per the identification key procedures by Gillies & Coetzee (refer SOP for identification of Anopheles mosquitoes by Gillies and Coetzee).

#### 6.2 Procedure for dissection

The investigators conducting dissection should be blinded from the treatment. Individual gravid female Anopheles mosquitoes will be dissected by gently pulling out the last two segments of the abdomen under a stereoscopic microscope at 0.7x magnification as described previously on the SOP MiS004 and procedures by Detinova et al [1]. Follicular ovaries and eggs development stages will be observed under a stereoscopic microscope. Detailed procedures for ovarian dissection of female mosquitoes are as follow;

1. Prepare and mark the dissecting slide.
2. Record the status and age of the mosquitoes at dissection in a dissection form. All mosquitoes at 72hrs post-collection should be gravid at the time of dissection.
3. Mount a freshly killed gravid female mosquito on either its left or back side on a slide with its abdomen pointing to the right
4. Use a dissecting needle on the thorax to hold the mosquito stationary while separate the abdomen from the head and thorax
5. Add a drop of distilled water on the last two segments, 6^th^ and 7^th^ sternites of the mosquito abdomen (figure 1)
6. Place the slide under a stereoscopic dissecting microscope (at a low magnification power 0.7x), using a dark background
7. Gently pulling off the last two segments of the mosquito abdomen using a dissecting needle on the right
8. Use the needle to separate the ovaries from other internal material. **For better visualization of the ovarian follicles, it is important that while separating the ovaries to cut on the common oviduct instead of separating from lateral oviduct**
9. Wash off fat and other debris by rinsing the ovaries with distilled water
10. Leave the slide with dissected ovaries and eggs to air dry

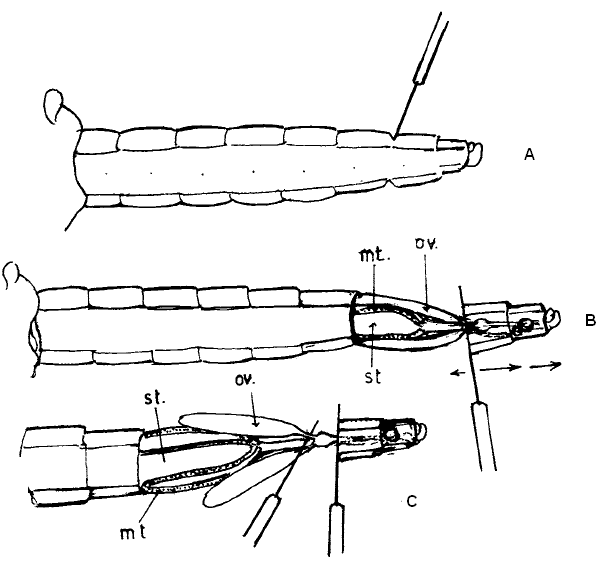

**Figure 1.** Extraction of ovaries. *ov = ovary, st = stomach, mt = malpigian tubules*

#### 6.3 Scoring Egg maturation and development

1. The standard Christopher scale will be used to score the development stage of the eggs (figure 2)

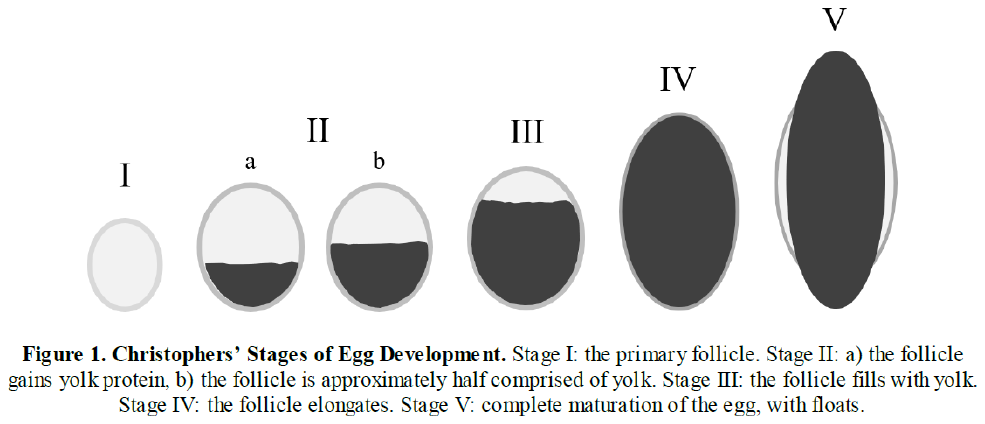

***Figure 2****: Showing Christopher’s phases of egg development in mosquito ovary [2]*

2. Observe the ovaries and eggs under the stereoscopic microscope. Sometimes you may need to add a small drop of water for better visualization.
3. Report the developmental status of ovarian follicles/eggs according to Christopher’s stage of egg development [2, 3] (I to IV = not undergone fully development (less elongated/spherical shape) to V (eggs have fully developed normal elongated, boat-shaped/sausage shape with floats).
4. Females Anopheles mosquitoes are categorized as **‘‘fertile’’** if the Christopher’s stage of egg development is “V” and **‘‘infertile’’** if stage I to IV. If eggs at different stages of development are observed, indicate this in the “egg stage” column of the data collection form. If both stage IV and stage V eggs are observed, record this as “inconclusive”.
5. Record the image showing developmental status of ovaries and eggs (Figure 2 and 3) in the mosquito dissection form**.** Take picture with a camera microscope and save the images into a tablet PC using a unique ID incorporating the test number, replicate number and mosquito number e.g T1R2M3. Please remember to rename each image corresponding to the label on the dissecting slide.
6. A Second readings should be done separately by a different investigator using the slide or on the picture if the second reading cannot be done on the same day.
7. In case of discrepancy a third reader should score the results.

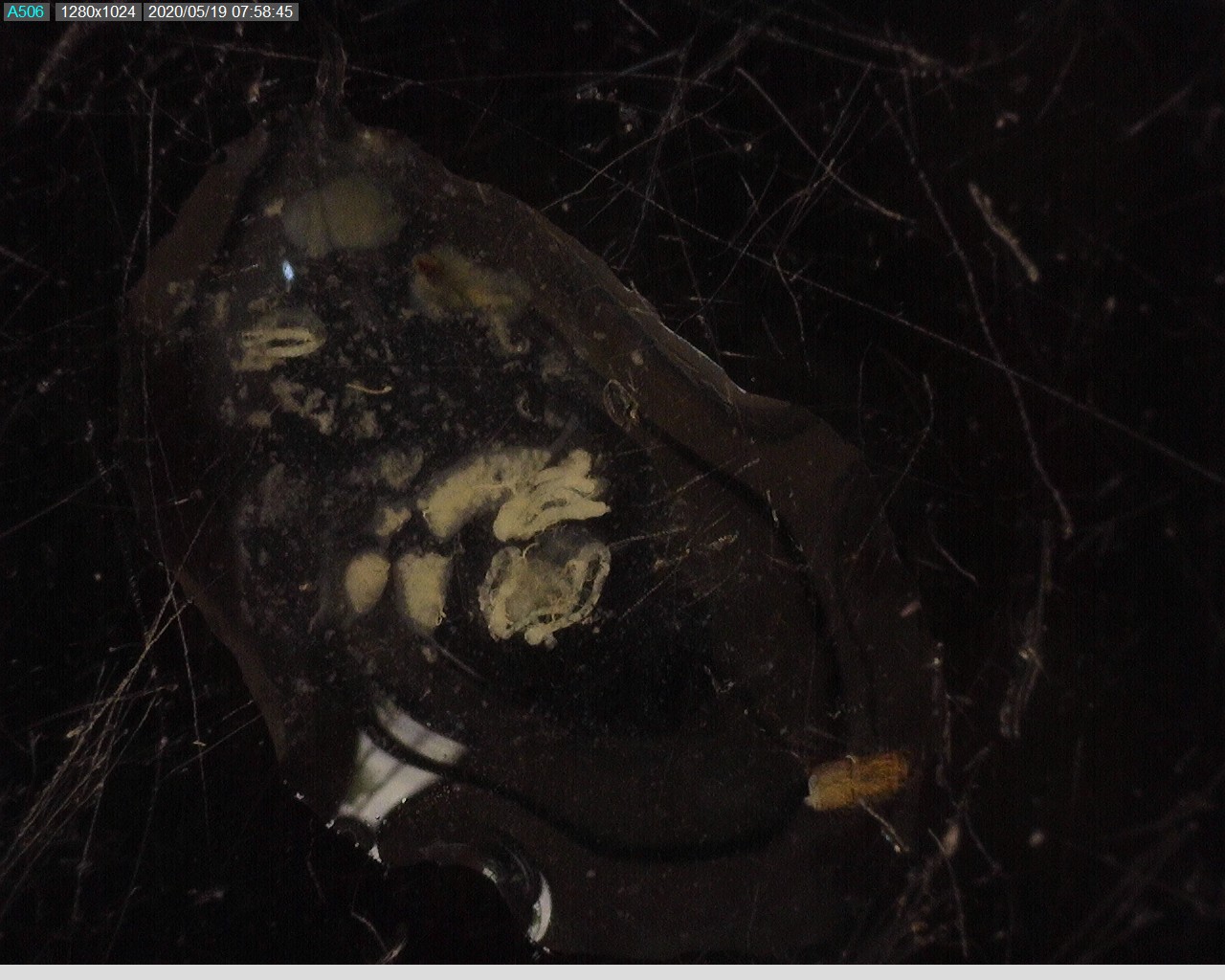

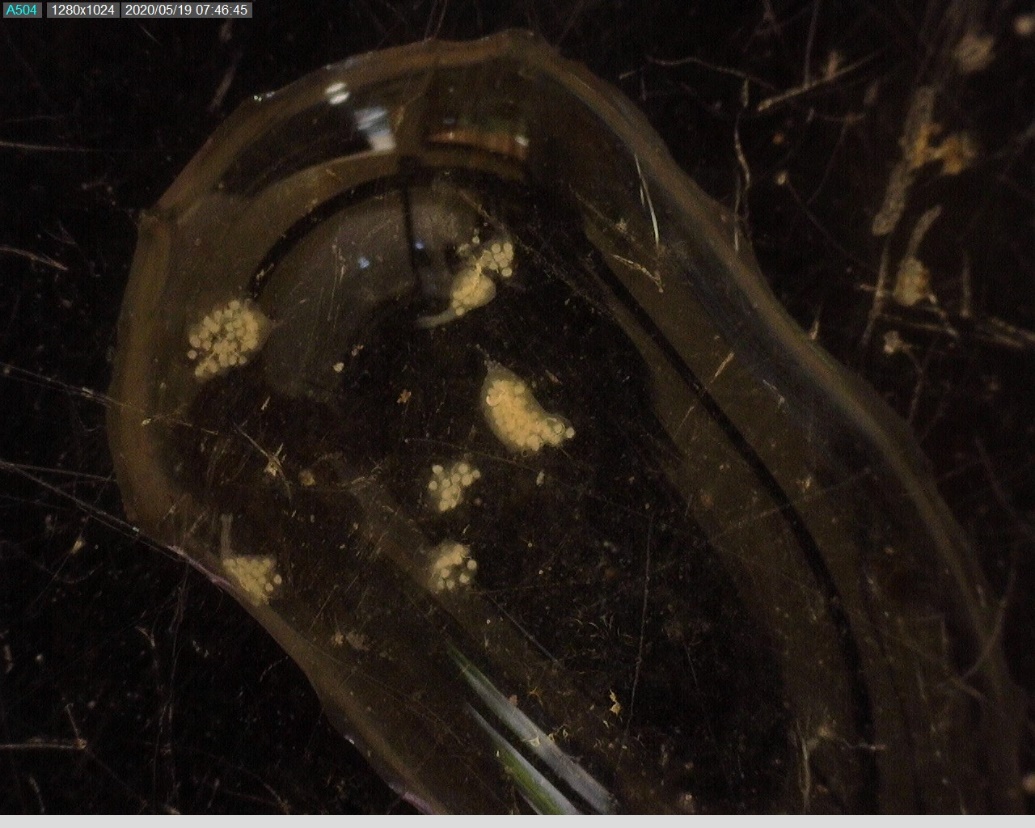

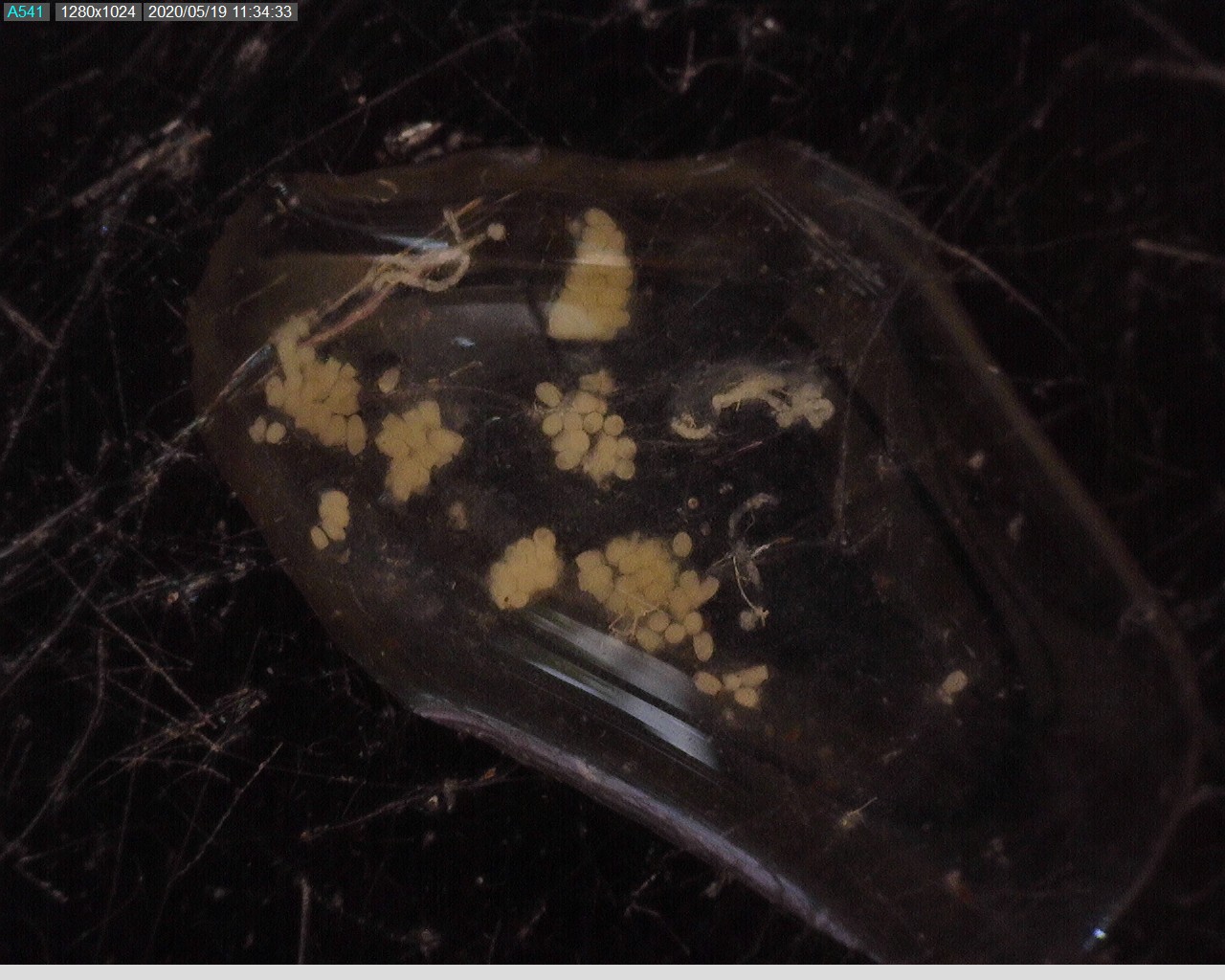

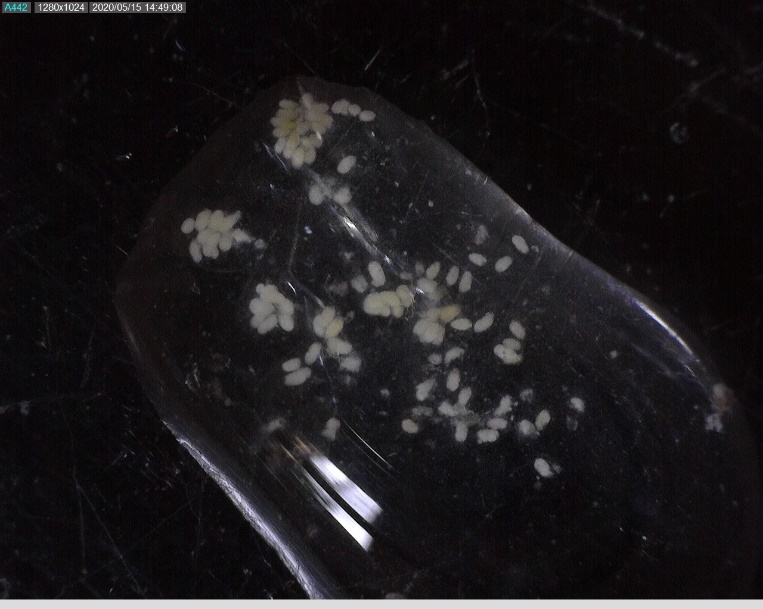

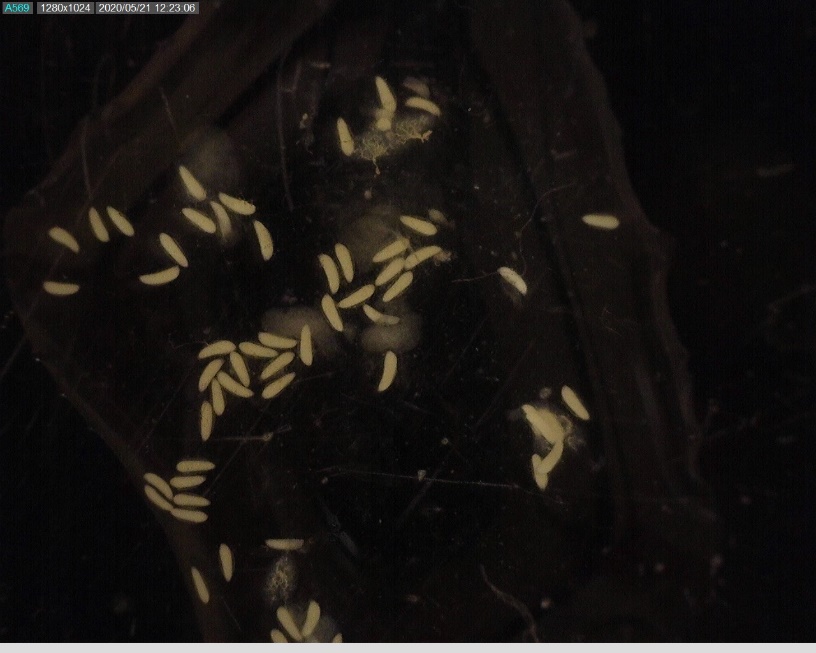

Figure 3

E

D

C

B

A

## 6.4

*Figure 3 shows the appearance of the eggs at different stages of development under a digital microscope (A. Stage I eggs, B. Stage II eggs, C. Stage III eggs D. Stage IV eggs E. Stage V eggs).*

**Appendix 1: Mosquito dissection form**

**Dissection Form for Fertility Study.	FORM MiF015**

| **Date**  **dd/mm/yy** | **Test No.** | **Net number** | **Position No.** | **Replicate No.** | **Holding period** | **Type of test (cone/tunnel)** | **Feeding Status^3^** | **Age at Dissection^4^** | **Status at Dissection^5^** | **Fertility Status^6^(F, IF)** | **Egg Stages^7^** | **FW**  **Initials** |
| --- | --- | --- | --- | --- | --- | --- | --- | --- | --- | --- | --- | --- |

**Feeding Status^3^**G=gravid, SG=semi-gravid, F=Blood fed, UF=unfed;**Age at Dissection^4^**24hrs, 48hrs, 72hrs post-blood meal**; Status at Dissection^5^;** A=Alive, D =Dead;**Fertility Status^6^;** F= Fertile; IF= Infertile; **Egg Development Stages^7^** (I, II, III, IV, and V)
