## Supplementary file 3 for "Durability of three types of dual active ingredient long-lasting insecticidal net compared to a pyrethroid-only LLIN in Tanzania: protocol for a prospective cohort study nested in a cluster randomized controlled trial"

****RANDOM LATIN SQUARE DO FILE (STATA 15.1) PREPARED BY JACKIE COOK & XXX (LONDON SCHOOL OF HYGIENE AND TROPICAL MEDICINE)**

clear all

**set the number of observations you need here- we've got 6 x 6 so 6 observations...

set obs 6

*This bit sets up the values in the latin square

***you can change the seed for each set of 6 days- just input a random number

set seed 1234

**using _n just results in counting the number of rows- we've got 6 observations- so get 1 to 6

gen newcode = _n

**this generates 6 random numbers between 0 and 1 (with lots of decimals)

gen rand_values = runiform()

**by sorting on the random values, this randomly mixes up our orignal observations

sort rand_values

*Generate the columns of a latin square

**the numbers represent the hut numbers (1,2,3,4,5,6)

**mod gets the remainder of dividing one number (_n) by another (6)**

**this is a way of generating hte numbers of 1 to 6 in each of our columns

gen v1 = 1+mod(_n,6)

gen v2 = 1+mod(_n+1,6)

gen v3 = 1+mod(_n+2,6)

gen v4 = 1+mod(_n+3,6)

gen v5 = 1+mod(_n+4,6)

gen v6 = 1+mod(_n+5,6)

***currently the column numbers are in order (1 2 3 4 5 6)- we need to mix them up**

**these generate local macros (named a through to f) which are equivalent to what is in the var newcode ///

***at the numbered positions

local a = newcode[1]

local b = newcode[2]

local c = newcode[3]

local d = newcode[4]

local e = newcode[5]

local f = newcode[6]

***this is then a loop which cycles through the numbers in the grid and replaces them with the macro above**

***i.e. where there is a 1 it is replaced with whatever is first in newcode etc***

forvalues i = 1/6 {

recode v`i' (1=`a') (2=`b') (3=`c') (4=`d') (5=`e') (6=`f')

}

drop newcode rand_values

*reorder the rows, then transpose (so that they are now the ocolumns), then reorder the rows again

gen rnumber_cols = runiform()

sort rnumber_cols

drop rnumber_cols

***this flips your data so your columns become rows***

xpose , clear

gen rnumber_rows = runiform()

sort rnumber_rows

drop rnumber_rows

br

****repeat this for each 6 days until you have enough for the full 6 weeks****

***so columns represent days, rows represent volunteers and the number in the cell is the hut number***
