## Supplementary file 4 for "Durability of three types of dual active ingredient long-lasting insecticidal net compared to a pyrethroid-only LLIN in Tanzania: protocol for a prospective cohort study nested in a cluster randomized controlled trial"

Net Study informed consent agreement

Introduction

Good morning. My name is __________. I work with PAMVERC Malaria Prevention Trial Missungwi. We work together with the Missungwi District Health office, the National Institute for Medical Research, the Kilimanjaro Christian Medical College, London School of Hygiene and Tropical Medicine.

General purpose of the survey

We would like to include you in a study to find information on quality and durability of the LLIN (net) you have been given earlier.

We would like to confirm how long the insecticide will last on the net. This is done by testing the nets in many households in several villages to find out how long they remain effective against malaria-transmitting mosquitoes. The strength of the netting material will be also looked.

Procedure

If you are willing to participate in the study:

- we expect you to not give away or sell the study net;
- you are free to stop using the nets at any time but we would expect you to let investigators know the reasons why you stopped using them during the follow up survey and to allow inspection of the nets;

Your household has been selected for repeated follow up (every 6 months for 3 years). We could take your net away for analysis. These nets will be replaced at that time. I would therefore like to have your consent to be interviewed; this will last about 20-30 minutes. During the interview, I will ask you some questions about your household, the status of the nets given to you or your family members and how you use your net and side effect if any. I will ask you to show the net to me, so I can assess its quality by counting holes in the net. I will not damage the net, and after the interview, I will return it.

Adverse effects, risks and participant protection

There is a remote possibility that you may get malaria even while using nets. This might be possible due to biting of mosquitoes outdoors or early in the night while your family was not sleeping under the net. Thus, if you suffer from fever, you should immediately approach the health staff available in your nearby health centre for treatment of possible malaria, as detailed below.

We are aware that pyrethroid insecticides are being used to treat the nets in the malaria control programme. Permethrin, the insecticide used on the nets, has been tested before and has not been found to have any undue adverse effects in most people at the dose found on the nets. Transitory tingling or runny nose has been recorded when nets are used for the first time when taken from its package. There is no cause for alarm as these effects pass within a day or two

Voluntariness and confidentiality

It is entirely your choice to take part in or not take part in this survey as I have just described it. If you agree to take part, you can also decide not to answer any of the questions that you do not want to.. If at any point in time during the study you take the decision not to participate any further, you are free to do so immediately and it will have no consequences, for example, your net will not be taken back from you. However we will want to ask you question to find out the reason why you decided to no longer use the net. We would also expect you to retain the net until our next visit so we can inspect its condition. Your individual information will be kept private.

Costs and compensation for participating in the study

You will not be asked to pay anything for you to participate in this study. The study will not reimburse you with any payment for taking part in the study.

The London School of Hygiene and Tropical Medicine is the Sponsor and hold insurance policies which apply to this study

Thank you very much for your time. Would you like to take part in this survey?

Consent section

- The study has been explained to me,
- I have been given the opportunity to ask questions concerning this study. Any such questions have been answered to my full satisfaction.
- I understand participation is voluntary and I may revoke this consent at any time without penalty or loss of benefits,
- I agree that the data generated from this study can be used in the future for other malaria related research. Yes |___|	No	|___|

I agree to take part.

**Name of guardian/parent.................................……….............. Signature/Thumb print ..............................**

**Name of the witness.....................................................Signature…………………….......................................**

**Name of interviewer……………………….............................Signature............................................................**

If you have any questions or clarification pertaining to this survey please feel free to ask the field workers or you may contact Study staff; Mr Eliud Lukole, PAMVERC, 0766240101; Dr Jackline Mosha, NIMR Mwnza, 0754404140; Dr Alphaxard Manjurano, 0756026661;

If you have any questions about your rights as a study patient, or if you think your child has been injured because of this study, please contact the Chairman of the National Health Research Ethics Committee (NatHREC) on 0222 121 400/390]

**PROJECT COPY**

**Cluster Number: ___ ___Household Number: ___ ___ ___ Date: __ __/__ __/__ __**

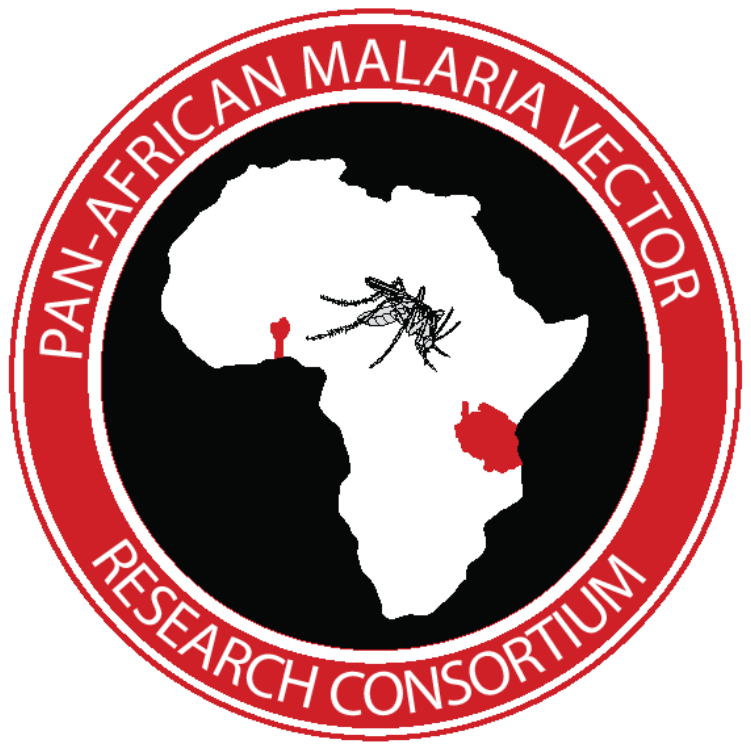
 Net Study Informed Consent agreement

Purpose of the survey

You received new LLIN and we would like to confirm how long the insecticide will last on the net. This is done by testing the nets in many households in several villages to find out how long they remain effective against malaria-transmitting mosquitoes. The strength of the netting material will be also looked.

Consent section

- The study has been explained to me,
- I have been given the opportunity to ask questions concerning this study. Any such questions have been answered to my full satisfaction.
- I understand participation is voluntary and I may revoke this consent at any time without penalty or loss of benefits,
- I agree that the data generated from this study can be used in the future for other malaria related research. Yes |___|	No	|___|

I agree to take part.

**Name of guardian/parent.................................……….............. Signature/Thumb print ..............................**

**Name of the witness.....................................................Signature…………………….......................................**

**Name of interviewer……………………….............................Signature............................................................**

If you have any questions or clarification pertaining to this survey please feel free to ask the field workers or you may contact Study staff; Mr Eliud Lukole, PAMVERC, 0766240101; Dr Jackline Mosha, NIMR Mwnza, 0754404140; Dr Alphaxard Manjurano, 0756026661;

If you have any questions about your rights as a study patient, or if you think your child has been injured because of this study, please contact the Chairman of the National Health Research Ethics Committee (NatHREC) on 0222 121 400/390]
