## Supplementary file 5 for "Durability of three types of dual active ingredient long-lasting insecticidal net compared to a pyrethroid-only LLIN in Tanzania: protocol for a prospective cohort study nested in a cluster randomized controlled trial"

**Volunteers information sheet and consent agreement (Hut trial)**

**V2.0: 17/02/2020**

Introduction

Good morning. My name is __________ . We work together with the Magu District Health office, the National Institute for Medical Research, the Kilimanjaro Christian Medical College, London School of Hygiene and Tropical Medicine.

Purpose of the survey

We would like to include you in a study to find out if different new nets products are effective against malaria. Malaria is transmitted by mosquitoes that carry the malaria parasite. The control interventions reduce the number of infected mosquitoes. We want to find out whether the LLINs reduce the number of mosquitoes flying in houses. It will provide information on which new LLINs works best to reduce mosquito numbers and malaria.

Participant Selection

We shall test the net in normal use in our huts. You and other volunteers will sleep under the net from 7 pm to 6 am from Monday to Friday. We can tell if the net is working by looking for dead mosquitoes in the hut in the morning. This evaluation will continue for seven weeks. We shall be regularly changing the nets. Sometimes the net will have been left unwashed, sometimes it will have been washed many times and sometimes nets will have been collected from the community. In this way we can work out if the nets are still working.

We are looking for local village volunteers to participate in the study. As a local you will have been exposed to biting by these mosquitoes. We are wanting to recruit adult males or females who are able to understand the purpose of the study. The persons have to be responsible and we would like them to help us collect the mosquitoes from the huts in the morning. We are looking for individuals who are able in principle to be available for the entire weeks of the study. You have to be willing to stay in the huts all night long. During the study you would be expected to sleep away from their families for a full 3 months. If there are more suitable volunteers than positions, we shall select individuals by lot.

Risks and Benefits:

By sleeping under a mosquito net you will obtain protection similar to what you have in your house if you use a insecticide treated net. But because in one of the huts you will also be sleeping under an untreated net we will monitor your fever every day. In the event that you contract malaria you must be aware of the signs and symptoms (which we shall tell you about), and inform us if you acquire any kind of fever so we can take other measures to treat you. If you have fever or suspected malaria a rapid diagnostic test will be taken for confirmation of parasites, and if positive you will be treated for free with an effective antimalarial drug.

You might also experience some reaction to the insecticide on the net, we request that you tell us about any sensations, side effects, or symptoms that may be due to sleeping under the treated net, so that we can take appropriate medical action. These symptoms may include headache, dizziness, sneezing, itching, tiredness. These side effects, if they occur at all, are temporary and are known to have no long-term consequence. A PAMVERC physician will be on hand to examine you. Any possible allergies to the insecticide formulation pointed out by the volunteers will be treated by the PAMVERC clinical team. You will receive a prophylaxis against malaria during the study.

Other possible discomforts include scratching caused by mosquito bites biting though the net, and blisters, redness or skin irritation caused by the insecticide itself.

Voluntariness and confidentiality

It is entirely your choice to participate in the activities I have just described it. You will not be penalized in any way if you refuse and will still receive all the services you currently receive if you choose not to participate. If at any point in time during the activities you take the decision not to participate any further, you are free to do so immediately, and it will have no consequences. Your individual information will be kept private. You will be given a code number and so any data is in reference to the code number rather than your name. This helps to protect your anonymity.

Costs and compensation for participating in the study

You will be paid a sum of 5000 Tz shilling a night for participating in the study as transport allowance.

Thank you very much for your time. Would you like to participate in the activities?

If you have any questions or clarification pertaining to this project please feel free to ask the field workers or you may contact Study staff; Ms. Jackline Martin, PAMVERC, 07574571391; Dr Jackline Mosha, NIMR Mwanza, 0754404140; Dr Alphaxard Manjurano, 0756026661;

If you have any question of this study, please contact the Chairman of the National Health Research Ethics Committee (NatHREC) on 0222 121 400/390.

**Volunteer COPY	Date: _____/______/________**

**Volunteers Informed Consent agreement**

Purpose of the survey

We are looking for volunteers to participate in a study to find out if different new nets products are effective against malaria. Malaria is transmitted by mosquitoes that carry the malaria parasite. We want to find out whether the new mosquito net reduce the number of mosquitoes entering in houses. It will provide information on which new LLINs works best to reduce mosquito numbers and malaria.

Consent section

- The study has been explained to me,
- I have been given the opportunity to ask questions concerning this study. Any such questions have been answered to my full satisfaction.
- I understand participation is voluntary and I may revoke this consent at any time without penalty or loss of benefits,
- I agree that the data generated from this study can be used in the future for other malaria related research. Yes |___|	No	|___|

I agree to take part.

**Name of participant.................................………........... Signature/Thumb print ..............................**

**Name of the witness..............................................Signature…………………….......................................**

**Name of interviewer………………………......................Signature............................................................**

If you have any questions or clarification pertaining to this project please feel free to ask the field workers and nurse or you may contact Study staff; Ms. Jackline Martin, PAMVERC, 07574571391; Dr Jackline Mosha, NIMR Mwanza, 0754404140; Dr Alphaxard Manjurano, 0756026661;

If you have any question of this study, please contact the Chairman of the National Health Research Ethics Committee (NatHREC) on 0222 121 400/390.

**PROJECT COPY**

**Cluster Number: ___ ___Household Number: ___ ___ ___ Date: __ __/__ __/__ __**

**Volunteers Informed Consent agreement**

Purpose of the survey

We are looking for volunteers to participate in a study to find out if different new nets products are effective against malaria. Malaria is transmitted by mosquitoes that carry the malaria parasite. We want to find out whether the new mosquito net reduce the number of mosquitoes entering in houses. It will provide information on which new LLINs works best to reduce mosquito numbers and malaria.

Consent section

- The study has been explained to me,
- I have been given the opportunity to ask questions concerning this study. Any such questions have been answered to my full satisfaction.
- I understand participation is voluntary and I may revoke this consent at any time without penalty or loss of benefits,
- I agree that the data generated from this study can be used in the future for other malaria related research. Yes |___|	No	|___|

I agree to take part.

**Name of guardian/parent.................................……….............. Signature/Thumb print ..............................**

**Name of the witness.....................................................Signature…………………….......................................**

**Name of interviewer……………………….............................Signature............................................................**

If you have any questions or clarification pertaining to this project please feel free to ask the field workers and nurse or you may contact Study staff; Ms. Jackline Martin, PAMVERC, 07574571391; Dr Jackline Mosha, NIMR Mwanza, 0754404140; Dr Alphaxard Manjurano, 0756026661; If you have any question of this study, please contact the Chairman of the National Health Research Ethics Committee (NatHREC) on 0222 121 400/390.
